# Disease and Participant-Related Correlates of Genetic Testing Completion for Hereditary Eye Disorders in a Cohort of Over 1400 Patients

**DOI:** 10.1101/2025.08.24.25334316

**Authors:** Dorothy T. Wang, Bani Antonio-Aguirre, Maria L. Ruggeri, Christy H. Smith, Kelsey S. Guthrie, Carolyn Applegate, Annabelle Pan, Setu P. Mehta, Kurt A. Dreger, Ishrat Ahmed, Jefferson J. Doyle, Mandeep S. Singh

## Abstract

**Objective:** To identify clinical and demographic predictors of genetic testing (GT) completion and diagnostic yield among patients with genetic eye disorders (GED) at a large U.S. tertiary academic center.

**Design:** Retrospective cohort study.

**Participants:** Patients with clinically diagnosed GEDs evaluated at the Wilmer Eye Institute’s Genetic Eye Disease (GEDi) Center between 2002 and 2025.

**Methods:** Demographic, clinical, and GT data were extracted. Bivariate analyses and multivariable logistic regression identified factors associated with GT completion and molecular diagnosis. Subgroup analyses examined racial disparities between Black, non-Hispanic White, and Other race participants.

**Main Outcome Measures:** Proportion of patients completing GT, molecular diagnostic yield, and clinical/demographic predictors of each.

**Results:** Of 1466 participants (median age at presentation 44 years, symptom onset 28 years; median follow-up 6 years), 74% (1088) completed genetic testing, with a likely molecular diagnosis achieved in 62%, inconclusive results in 22%, and no diagnosis in 16%. GT completion was more likely among younger participants with earlier symptom onset, and longer follow-up. Likely molecular diagnosis was more likely in participants with earlier symptom onset, worse visual acuity (VA), male sex, and syndromic or X-linked phenotypes. Black and Other race participants had significantly lower odds of completing GT (Black: OR [95% CI] 0.40 [0.30-0.53]; Other: 0.57 [0.36-0.92]) and receiving a likely molecular diagnosis (Black: 0.37 [0.26-0.51]; Other: 0.58 [0.35-0.98]), and consistently exhibited worse VA at both baseline and follow-up. Notably, among Black and Other race participants, disparities in GT completion and visual outcomes persisted despite equivalent or shorter time from presentation to GT completion, suggesting barriers arise independently of delays in care engagement. We identified 118 causative genes among participants with likely molecular diagnoses, with *ABCA4*, *USH2A*, *PRPH2*, *RHO*, and *BEST1* accounting for 47% of cases.

**Conclusions:** This is the largest single-center GED genetic testing cohort reported in the U.S. and reveals significant disparities in GT completion and yield by race, age, sex, and disease-level factors. Our findings underscore the need to expand early access to GT, diversify genomic databases, and address systemic barriers to ensure equity in GED diagnosis, clinical trial access, and delivery of emerging therapies.

## Introduction

Genetic eye disorders (GEDs), once considered untreatable, are now at the forefront of precision medicine due to advances in gene therapy, molecular diagnostics, and individualized care. The 2017 United States (U.S.) Food and Drug Administration (FDA) approval of voretigene neparvovec-rzyl (Luxturna) for *RPE65*-associated retinal dystrophy marked a paradigm shift in GED management and spurred investment in gene-based therapies.^1^ While gene-agnostic approaches are emerging, most approved and investigational GED treatments continue to rely on precise molecular diagnoses and detailed phenotypic characterization to determine eligibility and guide prognosis.^2^

Though individually rare, GEDs collectively affect approximately 1 in 1000 people worldwide, with inherited retinal diseases (IRDs) affecting approximately 1 in 3450.^3–5^ Over 400 genes have been implicated across syndromic and nonsyndromic forms, with inheritance patterns including autosomal recessive (most common), autosomal dominant, X-linked, and mitochondrial.^6^ This extensive genetic and phenotypic heterogeneity—further characterized by variable expressivity, penetrance, and overlapping clinical features—underscores the importance of accurate genotype-phenotype correlations for diagnosis, clinical trial inclusion, and therapeutic development.^7,8^

Numerous population-based GED studies have been conducted worldwide, spanning Europe,^9–20^ Asia,^21–30^ the Americas,^31–35^ Africa,^36–38^ and Oceania.^39,40^ The Foundation Fighting Blindness (FFB) IRD Gene Poll, the largest multinational IRD initiative to date, aggregated data from over 33,000 individuals across 41 clinical sites in 13 countries, providing a comprehensive global snapshot of IRD-associated genes and diagnoses.^41^ Several countries have advanced GED care through national registries, such as IRD-PT in Portugal, RD5000 in the Netherlands, and the Japan Eye Genetic Consortium, enabling systematic collection and sharing of clinical and genetic data.^9,42,43^ In contrast, the U.S. lacks a unified registry; most data derive from multicenter consortia like eyeGENE or disease-specific studies, limiting granular insight into genetic testing patterns and disparities at the individual center level.^34^ Few studies have characterized large, real-world GED cohorts within single tertiary care institutions in the U.S.^35^

Genetic testing is central to GED care, informing diagnosis, prognosis, family planning, and clinical trial eligibility. Understanding patterns of testing completion and barriers faced by those who do not pursue testing is essential to advancing equitable access to emerging therapies. Notably, even among those who undergo testing, results may be inconclusive due to limited panel sensitivity, uncertain variant classification, or phenotypic overlap with non-genetic conditions. However, factors associated with genetic testing completion remain poorly understood, which impedes the development of effective strategies to increase testing uptake and improve outcomes.

Recognizing this gap in knowledge, here we aimed to characterize the clinical and genetic features of GED patients evaluated at the Genetic Eye Diseases (GEDi) Center at the Wilmer Eye Institute, Johns Hopkins Hospital, a major referral center serving a large, demographically diverse GED population. Our study examines demographic and molecular patterns, genetic testing uptake, diagnostic yield, and potential disparities in access and outcomes, with the goal of informing more equitable delivery of precision eye care.

## Methods

### Ethics

This retrospective study was conducted at the Wilmer Eye Institute, Johns Hopkins Hospital, with approval from the Johns Hopkins University Institutional Review Board (IRB00213488). The study met criteria for a waiver of informed consent as a retrospective chart review. All study procedures adhered to the tenets of the Declaration of Helsinki and complied with Health Insurance Portability and Accountability Act (HIPAA) privacy regulations.

### Study Population and Data Collection

Patients evaluated in the Wilmer Eye Institute GEDi center between April 2002 and April 2025 for possible genetic etiology of an ocular condition were identified via electronic medical records (**Figure 1**).

**Figure 1.**
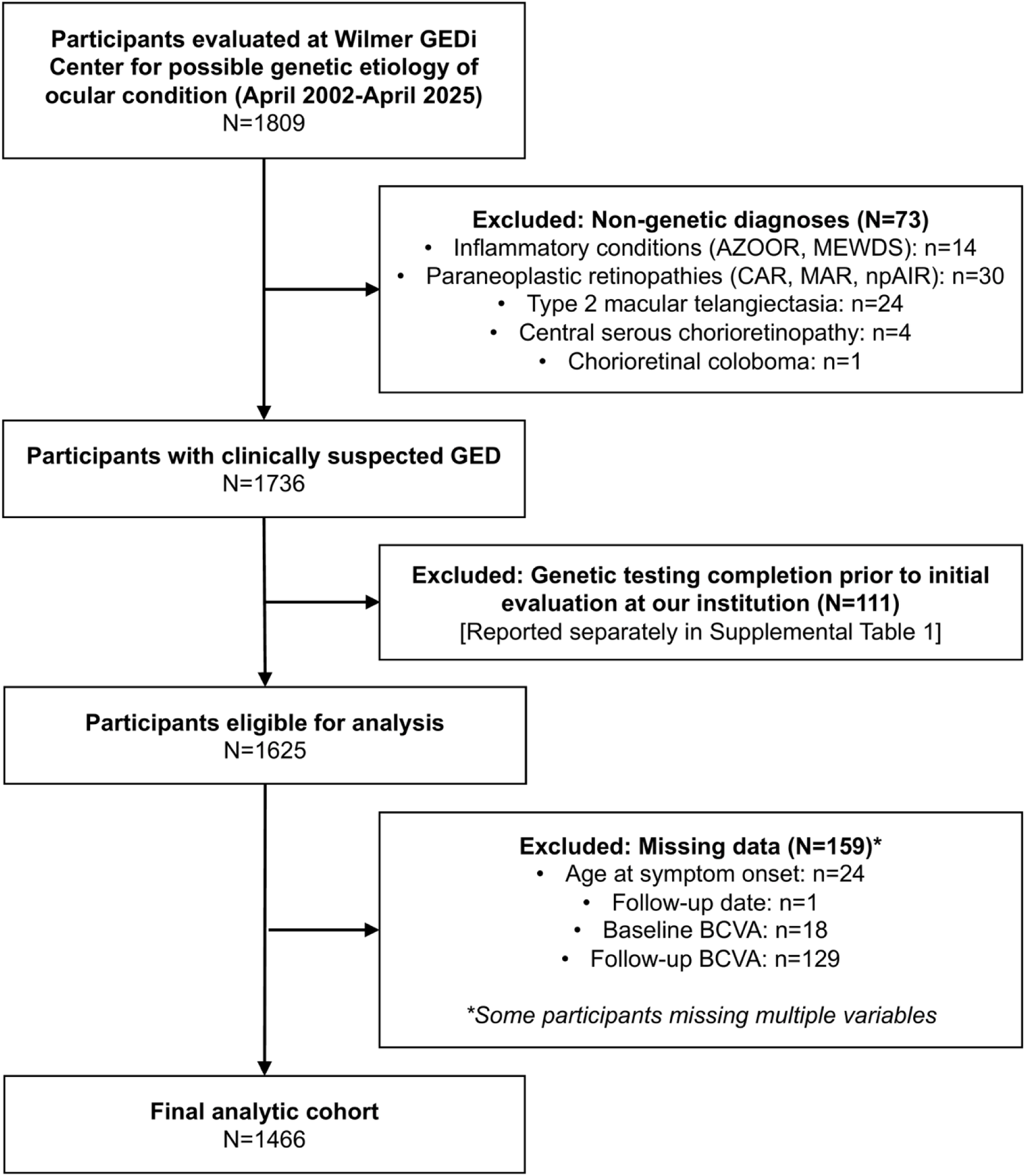
Flow diagram of patient selection. Of 1809 patients evaluated for possible genetic etiology of an ocular condition, 1466 met inclusion criteria for the final analytic cohort. Abbreviations: *GED* genetic eye disease; *AZOOR* acute zonal occult outer retinopathy; *MEWDS* multiple evanescent white dot syndrome; *npAIR* non-paraneoplastic autoimmune retinopathy; *CAR* cancer-associated retinopathy; *MAR* melanoma-associated retinopathy; *BCVA* best-corrected visual acuity.

Inclusion required clinical suspicion of a GED by a fellowship-trained GED specialist, based on clinical history, pedigree analysis, ophthalmic examination, and multimodal imaging (including fundus autofluorescence, optical coherence tomography, electroretinography, Goldmann visual field testing, and microperimetry, when available). Syndromic GEDs (e.g. Usher syndrome [USH], Bardet-Biedl, and Alstrom syndrome) required documented extraocular manifestations. The USH category was restricted to cases with both retinitis pigmentosa (RP) and sensorineural hearing loss; individuals with suspected autosomal recessive RP (arRP) or nonsyndromic retinal dystrophy alone were classified accordingly. Related individuals were included as independent observations.

Exclusion criteria included: (1) non-genetic etiologies (inflammatory conditions, paraneoplastic retinopathies, type 2 macular telangiectasia, central serous chorioretinopathy, chorioretinal coloboma); (2) genetic testing completed prior to initial evaluation at our institution (to ensure that baseline clinical characteristics and time-to-genetic-testing variables reflected assessments performed at our center); (3) insufficient clinical documentation to support a suspected GED diagnosis; and (4) incomplete demographic or clinical data. Participants excluded for prior genetic testing are reported separately (**Supplemental Table 1**).

While IRDs comprised the majority of diagnoses, we use the broader term “GED” to encompass inherited optic neuropathies, oculocutaneous albinism, corneoretinal dystrophies, and syndromic conditions with prominent ocular manifestations. A complete list of included clinical diagnoses is provided in **Supplemental Table 2**.

Collected demographic variables included sex, current age, race, and Hispanic/Latino ethnicity (HLE). Race was recorded according to National Institutes of Health classifications: American Indian or Alaska Native, Asian, Black or African American, Native Hawaiian or Other Pacific Islander, White, or Other.^44^ For predefined subgroup analyses, we compared participants in the Black, non-Hispanic White, or Other race categories; individuals identifying with other racial or ethnic groups were included in the overall analysis but excluded from these comparisons.

Extracted clinical data included the putative GED clinical diagnosis, age at symptom onset and presentation, symptom duration before presentation, follow-up duration, and best-corrected visual acuity (BCVA) at baseline and follow-up. BCVA was extracted from medical records in Snellen and subsequently converted to logarithm of the Minimum Angle of Resolution (logMAR) for analysis. The better-seeing eye was defined as the eye with the lower logMAR value, and the worse-seeing eye as the eye with the higher logMAR value; if both eyes had identical BCVA, the same value was assigned to both categories.

### Genetic Testing and Variant Classification

Genetic testing was performed as part of clinical care using GED panel tests ordered through Clinical Laboratory Improvement Amendments (CLIA)-certified laboratories; panel content varied over time depending on availability and clinical discretion. For participants who underwent genetic testing, we recorded genes and specific variants identified, time to genetic test completion, and suspected mode of inheritance as determined by provider documentation, family history, or molecular findings.

Variant classifications initially reflected designations provided by the testing laboratory according to American College of Medical Genetics and Genomics (ACMG)/Association for Molecular Pathology (AMP) guidelines.^45^ To incorporate updated evidence, all variants of uncertain significance (VUS) were systematically reviewed against current ClinVar annotations (accessed April 2026),^46^ and classifications were updated accordingly: VUS reclassified as pathogenic or likely pathogenic (P/LP) were incorporated into diagnostic classifications, and VUS reclassified as benign or likely benign (B/LB) were removed from consideration. Variants without updated ClinVar entries retained their original laboratory classifications.

Genetic test results were categorized as likely molecular diagnosis (positive), negative, or inconclusive according to the criteria outlined in **Table 1**. Classifications were determined by applying these criteria to reported variants in the context of the participant’s clinical phenotype and pedigree.

**Table 1.**
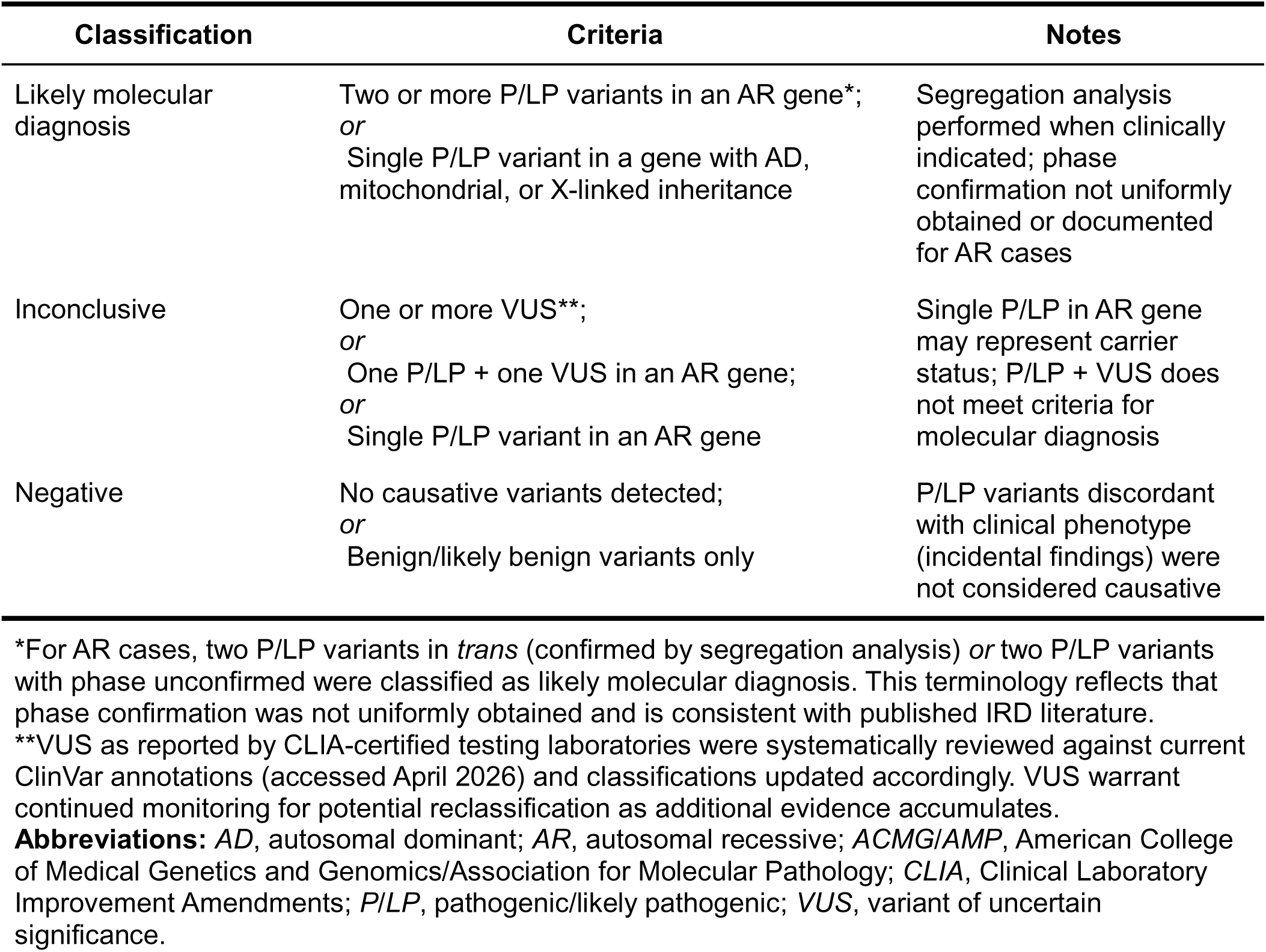
Genetic testing classification criteria.

| Classification | Criteria | Notes |
| --- | --- | --- |
| Likely molecular diagnosis | Two or more P/LP variants in an AR gene*;<br>*or*<br>Single P/LP variant in a gene with AD, mitochondrial, or X-linked inheritance | Segregation analysis performed when clinically indicated; phase confirmation not uniformly obtained or documented for AR cases |
| Inconclusive | One or more VUS**;<br>*or*<br>One P/LP + one VUS in an AR gene;<br>*or*<br>Single P/LP variant in an AR gene | Single P/LP in AR gene may represent carrier status; P/LP + VUS does not meet criteria for molecular diagnosis |
| Negative | No causative variants detected;<br>*or*<br>Benign/likely benign variants only | P/LP variants discordant with clinical phenotype (incidental findings) were not considered causative |

\*For AR cases, two P/LP variants in *trans* (confirmed by segregation analysis) *or* two P/LP variants with phase unconfirmed were classified as likely molecular diagnosis. This terminology reflects that phase confirmation was not uniformly obtained and is consistent with published IRD literature.
\*\*VUS as reported by CLIA-certified testing laboratories were systematically reviewed against current ClinVar annotations (accessed April 2026) and classifications updated accordingly. VUS warrant continued monitoring for potential reclassification as additional evidence accumulates.
**Abbreviations:** AD, autosomal dominant; AR, autosomal recessive; ACMG/AMP, American College of Medical Genetics and Genomics/Association for Molecular Pathology; CLIA, Clinical Laboratory Improvement Amendments; P/LP, pathogenic/likely pathogenic; VUS, variant of uncertain significance.

Likely molecular diagnosis was assigned if two or more P/LP variants were identified in an autosomal recessive gene, or if a single P/LP variant was found in a gene with autosomal dominant, mitochondrial, or X-linked inheritance. Segregation analysis was performed when clinically indicated and, when available, informed the final classification; however, segregation results were not systematically recorded in our database. Accordingly, we use the term “likely” molecular diagnosis” throughout this manuscript to reflect that phase confirmation was not uniformly obtained or documented for autosomal recessive cases with two P/LP variants. This terminology is consistent with published IRD literature, in which two P/LP variants identified in an autosomal recessive gene in a phenotypically concordant patient are variably described as “plausible”,^47^ “probable”,^48^ “likely”^49^ or “definitive”^19^ molecular diagnoses, even without formal phase confirmation.

Inconclusive results included cases with: (1) one or more VUS; (2) one P/LP variant and one VUS in an autosomal recessive gene; or (3) a single P/LP variant in an autosomal recessive gene. Negative results were assigned when no causative variants were detected or only B/LB variants were identified. P/LP variants discordant with the clinical phenotype were not considered causative and were classified as negative.

Causative genes were ranked by frequency among participants with likely molecular diagnoses.

### Statistical Analysis

De-identified data were analyzed using R version 4.5.1 (R Core Team, 2025).^55^ Descriptive statistics were generated for demographic, clinical, and genetic characteristics.

Bivariate analyses were conducted using chi-square or Fisher’s exact tests for categorical variables and Mann-Whitney U tests for nonparametric continuous variables. Analyses were stratified by (1) genetic testing status (completed genetic testing vs. untested), (2) diagnostic yield (likely molecular diagnosis vs. negative/inconclusive genetic test results), and (3) race/ethnicity (Black vs. non-Hispanic White; Other vs. non-Hispanic White). To account for multiple comparisons within each stratified analysis, we applied the Holm-Bonferroni correction, which produces adjusted p-values that control the family-wise error rate. Adjusted p-values <0.05 were considered statistically significant. Both raw and adjusted p-values are reported in Supplementary Tables; p-values reported in the main text represent adjusted values.

Multivariable logistic regression models were constructed to identify factors independently associated with (a) genetic testing completion and (b) diagnostic yield. Predictor selection was performed using least absolute shrinkage and selection operator (LASSO) regression with 10-fold cross-validation. For categorical predictors, reference groups were defined as the most prevalent category to ensure model stability and facilitate interpretability of effect estimates.

To assess the relationship between clinical phenotype and diagnostic yield, we performed univariable regression among participants who completed genetic testing. Diagnoses representing ≥2% of the tested cohort were included.

## Results

### Demographic, clinical, and genetic characteristics of the GED cohort

A total of 1809 patients were evaluated for possible genetic etiology of an ocular condition (**Figure 1**). After excluding non-genetic etiologies (n=73), patients who completed genetic testing prior to presentation (n=111; reported separately in **Supplemental Table 1**), and those with incomplete data (n=159), the final analytic cohort comprised 1466 participants with clinically suspected GEDs. The median age was 53 years (interquartile range [IQR]: 37-68; range: 3-99), with an approximately equal distribution of males and females. Most (62%; 902) participants self-identified as White, followed by Black or African American (24%; 350), with smaller proportions identifying as Asian (7%; 101), other races, or HLE (5%; 66). The median ages of symptom onset and presentation to the Wilmer Eye Institute were 28 (IQR 12-47) and 44 years (IQR 27-57), respectively. The median duration of symptoms prior to presentation was 6 years (IQR 1-20), and the median follow-up duration at Wilmer was 6 years (IQR 2-12). At baseline, median BCVA was approximately 20/40 Snellen in the better-seeing eye and 20/60 in the worse-seeing eye; at follow-up, values were approximately 20/50 and 20/100, respectively (**Table 2**).

**Table 2.** Demographic, clinical, and genetic characteristics of the genetic eye disease cohort (n=1,466). Data presented as median (IQR) or % (n).

| Parameter | Total (N=1466) |
| --- | --- |
| **Sex** |  |
| Male | 48% (704) |
| Female | 52% (762) |
| **Age** | 53 (IQR 37-68) years |
| **Race** |  |
| White | 62% (902) |
| Black or African American | 24% (350) |
| Native Hawaiian or Other Pacific Islander | <1% (2) |
| Asian | 7% (101) |
| American Indian or Alaska Native | <1% (5) |
| Other | 7% (106) |
| **Ethnicity** |  |
| Hispanic or Latino | 5% (66) |
| Not Hispanic or Latino | 95% (1400) |
| **Age of symptom onset** | 28 (IQR 12-47) years |
| **Age of presentation** | 44 (IQR 27-57) years |
| **Symptom duration prior to presentation** | 6 (IQR 1-20) years |
| **Duration of follow-up** | 6 (IQR 2-12) years |
| **Baseline BCVA, better-seeing eye** | 0.301 (IQR 0.097-0.699) |
| **Baseline BCVA, worse-seeing eye** | 0.477 (IQR 0.176-1) |
| **Follow-up BCVA, better-seeing eye** | 0.398 (IQR 0.097-0.903) |
| **Follow-up BCVA, worse-seeing eye** | 0.699 (IQR 0.301-1.301) |
| **Genetic testing status** |  |
| Completed genetic testing | 74% (1088) |
| Untested | 26% (378) |
| **Genetic testing result (n=1088)** |  |
| Likely molecular diagnosis | 62% (676) |
| Inconclusive | 22% (237) |
| Negative | 16% (175) |
| **Suspected mode of inheritance** |  |
| Autosomal Dominant | 9% (131) |
| Autosomal Recessive | 32% (465) |
| Autosomal Recessive/Dominant | 3% (47) |
| X-linked Recessive | 4% (59) |
| X-linked Dominant | <1% (1) |
| Mitochondrial | 1% (15) |
| Unknown after genetic testing | 25% (369) |
| Untested | 26% (379) |

Abbreviations: *IQR* = interquartile range; *BCVA* = best-corrected visual acuity (measured in logMAR)

Genetic testing was completed by 74 % (1088) of the cohort. Among those tested, 62% (676) had likely molecular diagnoses that explained the clinical phenotype, 22% (237) had inconclusive results, and 16% (175) had negative results. Across the full cohort, autosomal recessive inheritance was most frequently suspected (32%; 465), followed by autosomal dominant (9%; 131) and X-linked recessive (4%; 59); inheritance remained unknown in 51% (748) of cases, either due to lack of genetic testing completion or negative/inconclusive test results (**Table 2**). The median age at genetic testing was 46 years (IQR 29-61), and median duration of symptoms prior to genetic testing completion was 11 years (IQR 4-23). **Supplemental Figure 1** illustrates the distribution of age at genetic testing by mode of inheritance.

### Factors associated with genetic testing completion

Participants who completed genetic testing (n=1088) were significantly younger than those who did not (n=378; median age 50 vs. 60 years, adjusted p<0.001) and had earlier symptom onset (median 25 vs. 37 years, adjusted p<0.001) and age at presentation (41 vs. 49 years, adjusted p<0.001). Genetic testing completion differed significantly by race: among White (704/902) and Asian (84/101) participants, approximately 80% completed genetic testing, compared to 63% (219/350) of Black or African American participants (**Figure 2**, **Supplemental Table 3**).

**Figure 2.**
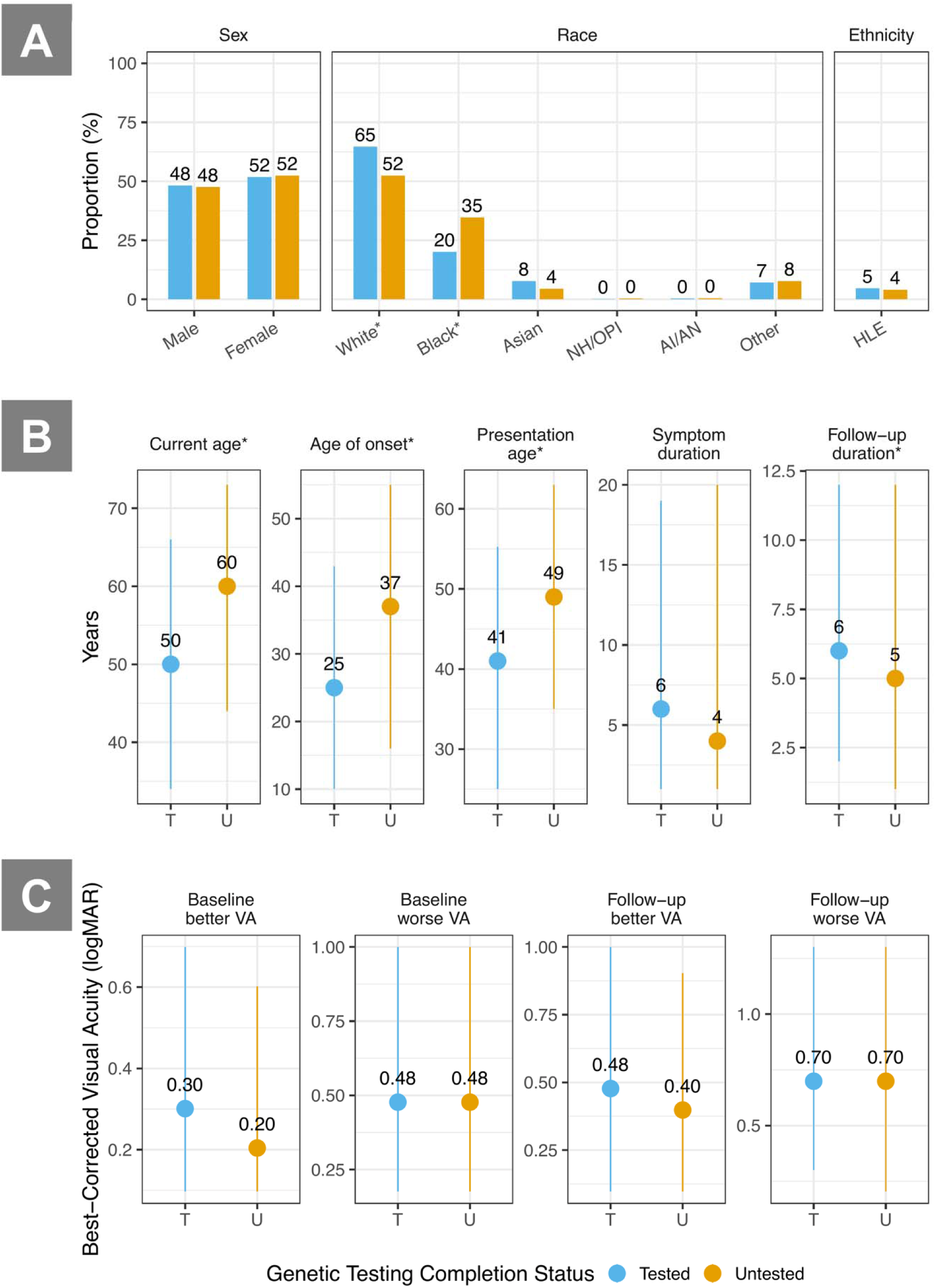
Demographic and clinical characteristics by genetic testing completion status. Asterisks indicate statistically significant differences (p<0.05). A) Proportions by sex, race, and ethnicity. B) Key age and duration measures by subgroup: current age, age at symptom onset, age at presentation, symptom duration prior to presentation, and duration of follow-up (all in years; circles indicate medians and lines indicate interquartile ranges). C) Best-corrected visual acuity (logMAR) for the better-seeing and worse-seeing eyes at baseline and follow-up. Abbreviations: *AI/AN* American Indian or Alaska Native; *NH/OPI* Native Hawaiian or Other Pacific Islander; *HLE* Hispanic or Latino ethnicity; *VA* visual acuity; *T* tested (completed genetic testing); *U* untested

Genetic testing rates also varied by clinical diagnosis. Among diagnostic categories representing at least 1% of the cohort, the highest completion rates were observed in choroideremia (100%), USH (90%), achromatopsia (86%), STGD (86%), Leber Congenital Amaurosis/Severe early-onset retinal dystrophy (LCA/SEORD; 82%), cone-rod dystrophy (CRD; 81%), and hereditary optic neuropathy (HON; 76%). The lowest rates were observed in pattern dystrophy (PD) (44%), L-ORD (50%), and unspecified macular dystrophy (56%), all of which are predominantly adult-onset conditions with significant clinical overlap with other retinal diseases or limited perceived progression. A full breakdown of diagnoses—including associated ages of symptom onset/presentation, genetic testing status, and results—is provided in **Supplemental Table 2.** Follow-up duration was longer in the tested group (6 vs. 5 years, adjusted p<0.05). (**Figure 2, Supplemental Table 3**).

In multivariable logistic regression, Black or African American race (odds ratio [OR] 0.40; 95% confidence interval [CI] 0.30-0.53; p<0.001) and Other race (OR 0.57; 95% CI 0.36-0.92; p<0.05) were associated with lower odds of genetic testing completion compared to White race. Later age of symptom onset (70+ years) was associated with lower odds of genetic testing completion (OR 0.23; 95% CI 0.12-0.43; p<0.001) compared to the reference group (10-19 years), while early symptom onset (0-9 years) showed a trend toward higher odds of genetic testing completion that approached but did not reach statistical significance (OR 1.53; 95% CI 1-2.37; p=0.05). Compared to RP, participants diagnosed with STGD had higher odds of genetic testing completion (OR 2.00; 95% CI 1.32-3.08; p=0.001), whereas those diagnosed with PD had lower odds (OR 0.35; 95% CI 0.21-0.57; p<0.001) (**Figure 3, Supplemental Table 4**).

**Figure 3.**
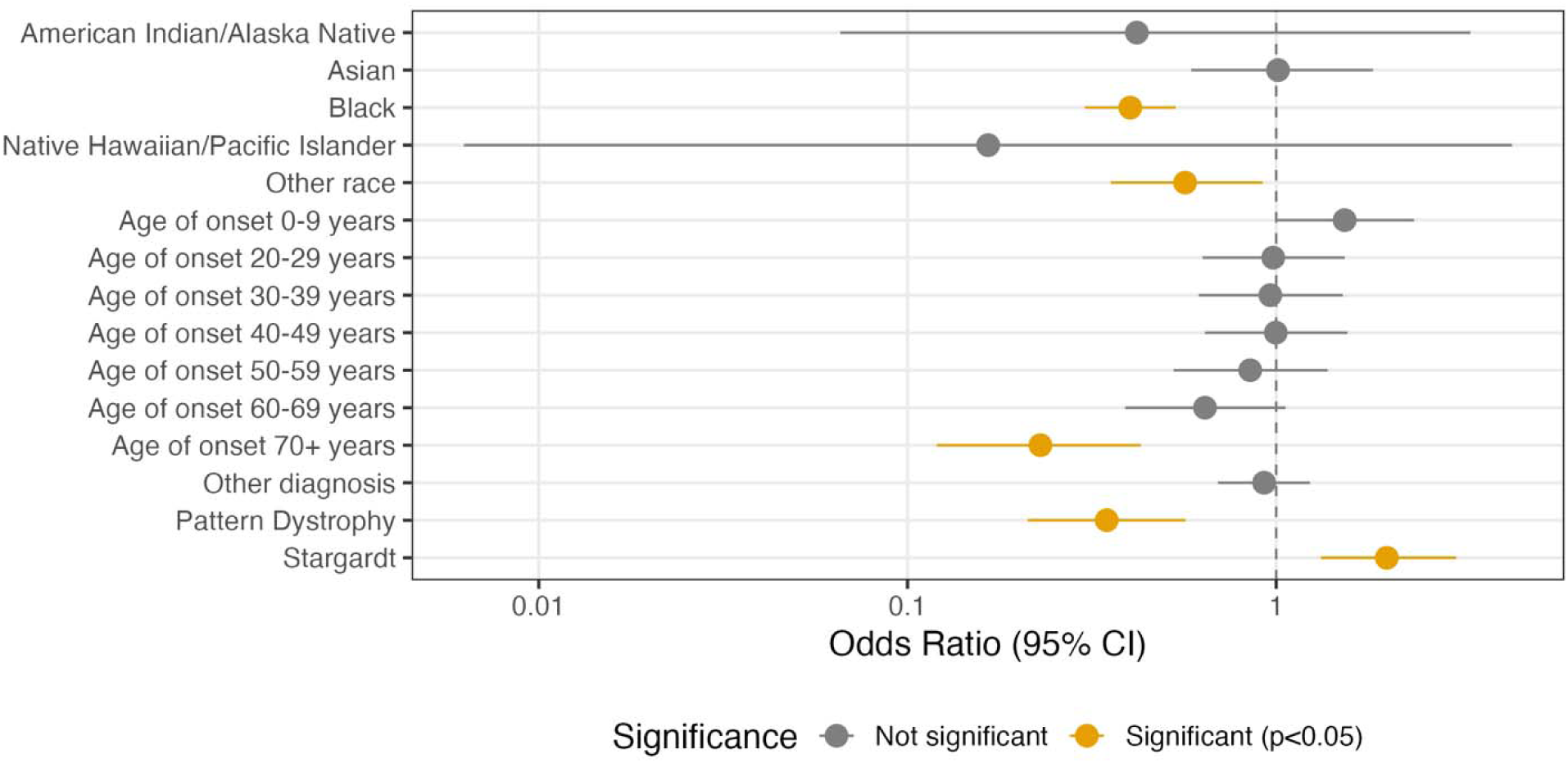
Multivariable logistic regression of factors associated with genetic testing completion. Predictors were selected using LASSO regression and included in a logistic regression model (n=1466). The outcome was defined as completion of genetic testing (tested vs. untested). Reference groups were based on the most frequent category for each variable (White race, age of symptom onset 10-19 years. Odds ratios (ORs) are shown as dots; horizontal lines represent 95% confidence intervals (CIs). CIs that do not cross 1 indicate statistical significance (p<0.05); exact p-values are listed in **Supplemental Table 4.** Model fit was assessed using multiple metrics (AUC=0.69, Hosmer-Lemeshow goodness-of-fit test p=0.92, generalized VIF ranged from 1.01 to 1.04 indicating no multicollinearity among predictors). Abbreviations: *LASSO* least absolute shrinkage and selection operator; *OR* odds ratio; *CI* confidence interval; *AUC* area under the curve; *VIF* variance inflation factor

### Factors associated with molecular diagnosis among participants who completed genetic testing

Among 1088 participants who completed genetic testing, factors associated with diagnostic yield were evaluated by comparing those with likely molecular diagnoses (n=676) versus negative or inconclusive (n=412) results. Male sex had higher diagnostic yield (67% [349/524] vs. 58% [327/564]; adjusted p<0.05). Younger age (median 46 vs. 56.5 years), earlier symptom onset (median 20 vs. 36 years), and younger age at presentation (median 37 vs. 48 years) were all associated with increased likelihood of a likely molecular diagnosis (adjusted p<0.001 for all). Longer symptom duration prior to presentation (8 vs. 4 years; adjusted p<0.001) and prior to genetic testing completion (13 vs. 9 years; adjusted p<0.01) were both associated with higher likelihood of a likely molecular diagnosis (**Table 3**).

**Table 3.** Factors associated with molecular diagnosis among participants who completed genetic testing (n=1,088). Data presented as % (n) or median (IQR).

| Parameter | Likely molecular diagnosis (N=676) | Negative or inconclusive test results (N=412) | Raw p-value | Adjusted p-value |
| --- | --- | --- | --- | --- |
| **Sex** | | | $\chi^2$ p=0.004 | 0.041* |
| Male | 52% (349) | 42% (175) |  |  |
| Female | 48% (327) | 58% (237) |  |  |
| **Age** | 46 (IQR 31-63) years | 56.5 (IQR 38-69) years | M-W p=2.7e-07 | 4.2e-06* |
| **Race** |  |  |  |  |
| White | 70% (471) | 57% (233) | $\chi^2$ p=1.9e-05 | 2.7e-04* |
| Black or African American | 15% (100) | 29% (119) | $\chi^2$ p=2.9e-08 | 5.0e-07* |
| Asian | 8% (55) | 7% (29) | $\chi^2$ p=0.411 | 1 |
| Native Hawaiian or Other Pacific Islander | <1% (1) | 0% (0) | Fisher's p=1 | 1 |
| American Indian or Alaska Native | <1% (2) | <1% (1) | Fisher's p=1 | 1 |
| Other | 7% (47) | 7% (30) | $\chi^2$ p=0.801 | 1 |
| **Ethnicity** | | | $\chi^2$ p=0.725 | 1 |
| Hispanic or Latino | 4% (30) | 5% (21) |  |  |
| Not Hispanic or Latino | 96% (646) | 95% (391) |  |  |
| **Age of symptom onset** | 20 (IQR 9-35) years | 36 (IQR 15-55) years | M-W p=1.4e-17 | 2.7e-16* |
| **Age at presentation** | 37 (IQR 21.8-52) years | 48 (IQR 30-59) years | M-W p=7.4e-09 | 1.3e-07* |
| **Symptom duration prior to presentation** | 8 (IQR 2-22) years | 4 (IQR 1-14) years | M-W p=1.4e-06 | 2.1e-05* |
| **Symptom duration prior to GT completion** | 13 (IQR 5-27) years | 9 (IQR 3-18) years | M-W p=8.4e-05 | 0.001* |
| **Duration of follow-up** | 6 (IQR 3-12) years | 6 (IQR 2-11) years | M-W p=0.041 | 0.33 |
| **Baseline BCVA, better-seeing eye** | 0.301 (IQR 0.097-0.699) | 0.204 (IQR 0.073-0.559) | M-W p=0.003 | 0.038* |
| **Baseline BCVA, worse-seeing eye** | 0.498 (IQR 0.204-1) | 0.398 (IQR 0.176-0.927) | M-W p=0.071 | 0.43 |
| **Follow-up BCVA, better-seeing eye** | 0.505 (IQR 0.176-1) | 0.398 (IQR 0.097-0.875) | M-W p=2.8e-04 | 0.003* |
| **Follow-up BCVA, worse-seeing eye** | 0.796 (IQR 0.301-1.301) | 0.602 (IQR 0.301-1.176) | M-W p=0.004 | 0.041* |
| **Suspected mode of inheritance\*\*** |  |  | Fisher's p=0.046 | 0.33 |
| Autosomal Dominant | 19% (124) | 14% (7) |  |  |
| Autosomal Recessive | 64% (426) | 78% (39) |  |  |
| X-linked Recessive | 9% (58) | 2% (1) |  |  |
| Autosomal Recessive/Dominant | 7% (46) | 2% (1) |  |  |
| Mitochondrial | 2% (14) | 2% (1) |  |  |
| X-linked Dominant | <1% (1) | 0% (0) |  |  |

\*Adjusted p<0.05 (Holm-Bonferroni correction for multiple comparisons)
\*\*N=369 participants with unknown inheritance patterns (due to inconclusive/negative genetic testing results) were excluded from inheritance-related comparisons.
Abbreviations: BCVA = best-corrected visual acuity (logMAR); GT = genetic testing; IQR = interquartile range; M-W = Mann-Whitney U test; $\chi^2$ = chi-square test; Fisher's = Fisher's

| Parameter | Likely molecular diagnosis (N=676) | Negative or inconclusive test results (N=412) | Raw p-value | Adjusted p-value |
| --- | --- | --- | --- | --- |

Diagnostic yield varied by race: 67% (471/704) of White, 46% (100/219) of Black or African American, 65% (55/84) of Asian, and 61% (47/77) of Other race participants were classified as likely molecular diagnoses. Likely molecular diagnoses were also observed in 67% (2/3) of American Indian or Alaska Native participants, and in the single Native Hawaiian or Other Pacific Islander participant (**Table 3**). Baseline visual function in the better-seeing eye was worse among participants with likely molecular diagnoses compared to those with negative/inconclusive results (median 0.301 vs. 0.204 logMAR; adjusted p<0.05), as was follow-up BCVA in both eyes (adjusted p<0.05 for both) (**Table 3**).

In multivariable regression, male sex was associated with increased odds of a likely molecular diagnosis (OR 1.37; 95% CI 1.05-1.78; p<0.05). Compared to White participants, Black participants had significantly lower odds of a likely molecular diagnosis (OR 0.37; 95% CI 0.26-0.51; p<0.001), as did participants of Other race (OR 0.58; 95% CI 0.35-0.98; p<0.05). Later age at symptom onset was associated with progressively lower odds of a likely molecular diagnosis, with a clear stepwise decline across older age groups relative to the reference group (ages 0-9): 40-49 (OR 0.52; 95% CI 0.33-0.82; p<0.01), 50-59 (OR 0.20; 95% CI 0.12-0.33), 60-69 (OR 0.20; 95% CI 0.11-0.35), and 70+ years (OR 0.12; 95% CI 0.04-0.32) (all p<0.001 unless otherwise indicated). Finally, worse follow-up BCVA in the better-seeing eye was associated with increased odds of a likely molecular diagnosis (OR 1.33 per 1.0 logMAR increase; 95% CI 1.08-1.66; p<0.01), suggesting that more severe visual impairment may be linked to genetically identifiable disease (**Figure 4, Supplemental Table 5**).

**Figure 4.**
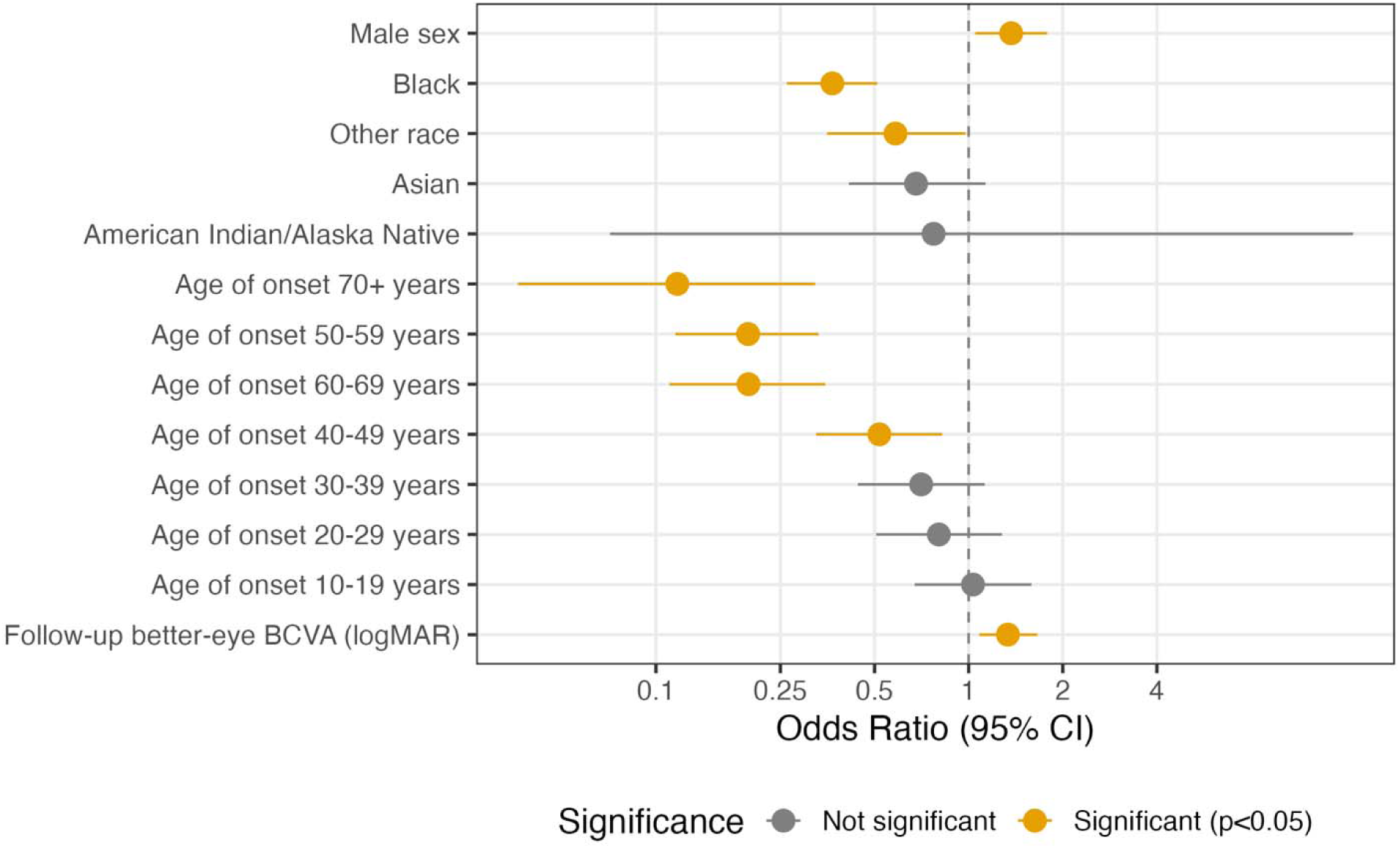
Multivariable logistic regression of factors associated with molecular diagnostic yield among participants who completed genetic testing. Predictors were selected using LASSO regression and included in a logistic regression model contrasting genetic test results of likely molecular diagnoses vs. negative or inconclusive results. Odds ratios (ORs) are shown as dots; horizontal lines represent 95% confidence intervals (CIs). Reference groups for categorical variables were set as the most frequent category (Female sex, White race, age at onset 0-9 years). Exact p-values are listed in **Supplemental Table 5**. Model fit was assessed using multiple metrics (AUC=0.71, Hosmer-Lemeshow goodness-of-fit test p=0.79, generalized VIF ranged from 1.00 to 1.03 indicating no multicollinearity among predictors). Note: Native Hawaiian/Other Pacific Islander race excluded due to small sample size (n=1) and unstable estimates, thus a total of n=1086 were included in the model. Abbreviations: *LASSO* least absolute shrinkage and selection operator; *BCVA* best-corrected visual acuity; *logMAR* logarithm of the Minimal Angle of Resolution; *OR* odds ratio; *CI* confidence interval

### Spectrum of clinical diagnoses and genetic etiologies in the GED cohort

The most common putative clinical diagnoses were RP (32.3%), STGD (17.4%), PD (7.2%), cone dystrophy (CD; 6.9%), CRD (6.8%), USH (4.6%), HON (2.9%), unspecified macular dystrophy (2.7%), Best disease (e.g. Bestrophinopathy & Best vitelliform macular dystrophy; BEST; 2.3%), and oculocutaneous albinism (OCA; 1.7%), together accounting for 85% (1240 of 1466) of the cohort. **Figure 5** depicts genetic testing completion rates and positive diagnostic yields for diagnostic categories representing at least 1% of the cohort. Diagnostic yield varied substantially by phenotype, with the highest rates in USH (93%; 56/60) and BEST (91%; 21/23), followed by choroideremia (89%; 16/18), STGD (83%; 182/220), and retinoschisis (77%; 10/13). Lower yields were observed in RP (58%; 207/354), OCA (53%; 8/15), CRD (49%; 39/80), HON (41%; 13/32), PD (34%; 16/47), CD (28%; 18/65), L-ORD (25%; 3/12), and unspecified macular (18%) and retinal (15%) dystrophies **(Supplemental Table 2, Figure 5)**.

**Figure 5.**
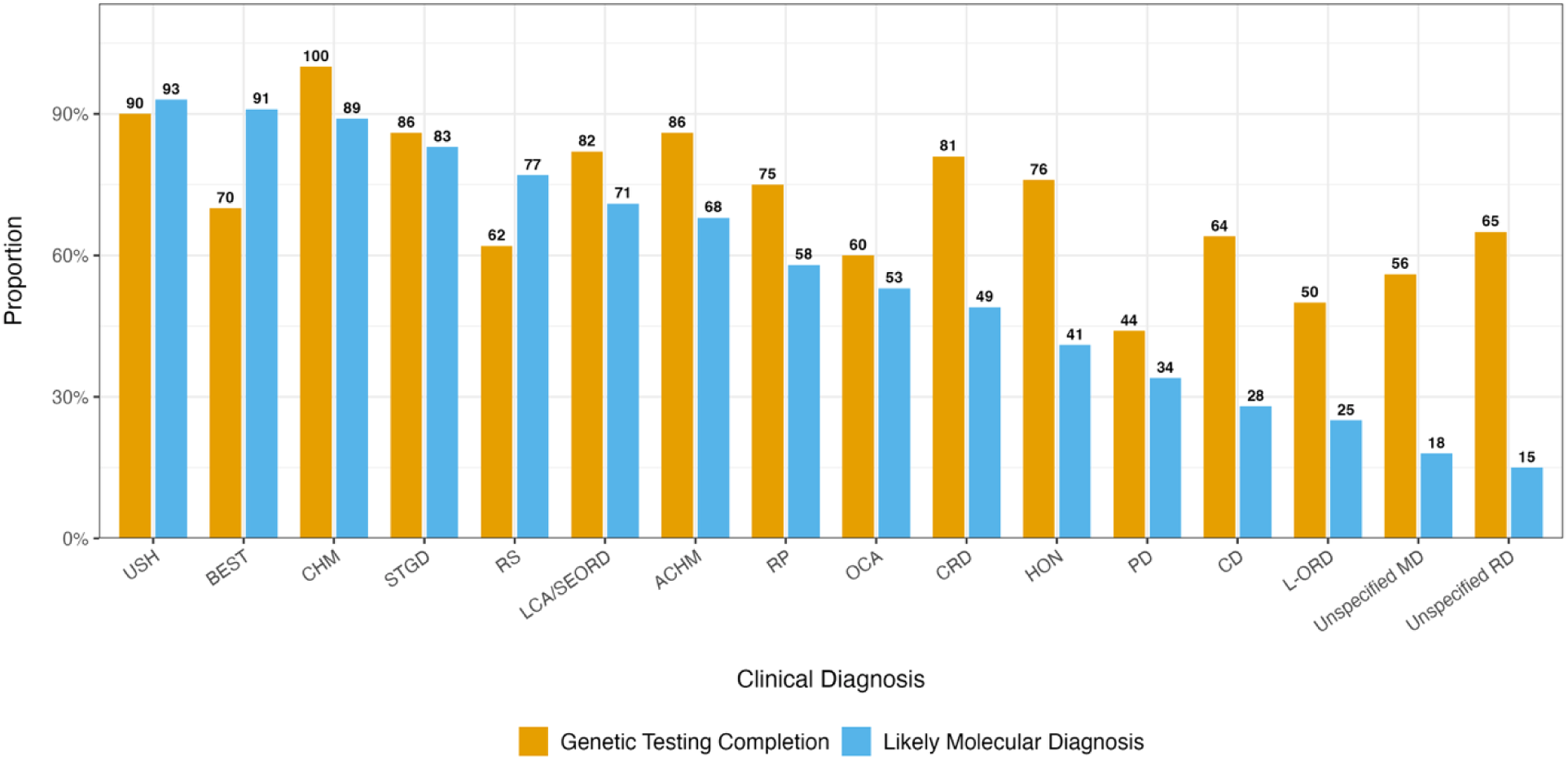
Likely molecular diagnosis and genetic testing completion rates among inherited eye disease participants, grouped by common putative clinical diagnoses. Blue bars indicate the proportion of patients achieving a likely molecular diagnosis in each phenotype category; orange bars indicate the genetic testing completion rate. Only clinical diagnoses representing ≥1% of the total cohort are shown. Abbreviations: *USH* Usher syndrome; *BEST* Best disease; *CHM* choroideremia; *STGD* Stargardt disease; *RS* retinoschisis; *LCA/SEORD* Leber Congenital Amaurosis/Severe early-onset retinal dystrophy *ACHM* achromatopsia; *RP* retinitis pigmentosa; *OCA* oculocutaneous albinism; *CRD* cone-rod dystrophy; *PD* pattern dystrophy; *HON* hereditary optic neuropathy; *CD* cone dystrophy; *L-ORD* late-onset retinal dystrophy; *MD* macular dystrophy; *RD* retinal dystrophy

Univariable logistic regression (**Figure 6)** confirmed that STGD (OR 3.40, 95% CI 2.28-5.17) and USH (OR 9.94, 95% CI 3.97-33.3) were associated with higher odds of a likely molecular diagnosis compared to RP (p<0.001), whereas CD (OR 0.27, 95% CI 0.15-0.48, p<0.001) and PD (OR 0.37, 95% CI 0.19-0.69, p<0.01) were associated with significantly lower odds.

**Figure 6.**
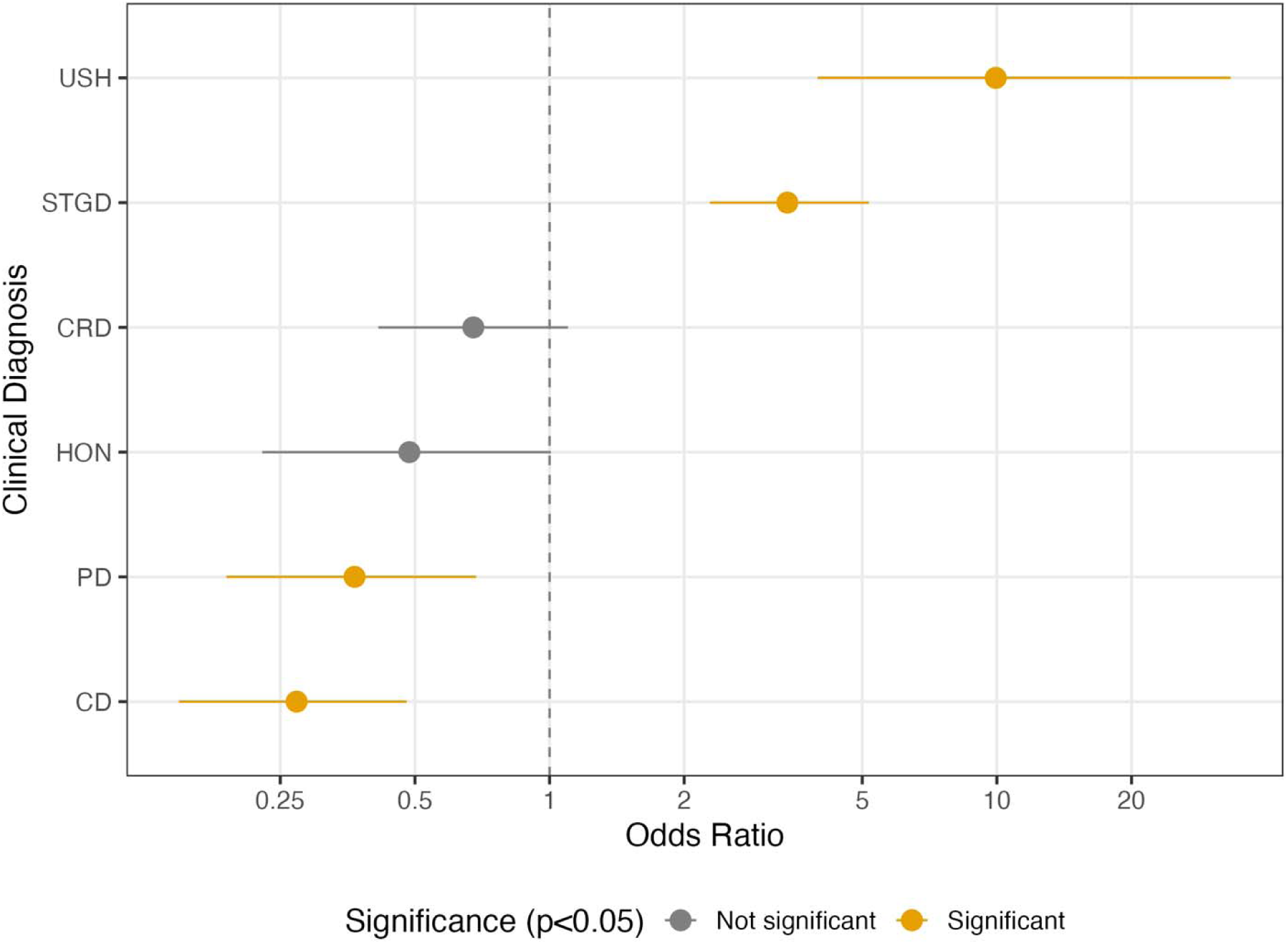
Likelihood of molecular diagnosis by common putative clinical diagnoses. Odds ratios and 95% confidence intervals are shown from univariate logistic regression of clinical diagnostic groups, with RP as the reference group (not shown). Significant associations (p<0.05) are shown in orange. Clinical diagnoses representing ≥2% of the total cohort are included. Abbreviations: *RP* retinitis pigmentosa; *USH* Usher syndrome; *STGD* Stargardt disease; *CRD* cone-rod dystrophy; *PD* pattern dystrophy; *HON* hereditary optic neuropathy; *CD* cone dystrophy

Among 673 participants with a molecular diagnosis and complete variant data, the most common disease-causing genes were *ABCA4* (26.2%), *USH2A* (9.5%), *PRPH2* (4.7%), *RHO* (4.0%), and *BEST1* (3.1%), together accounting for 47% (319/676) of cases with likely molecular diagnoses (**Table 4**). In total, 118 distinct disease-associated genes were identified (**Figure 7, Supplemental Table 6**), with 11 genes found in at least 10 participants and 28 found in at least five. The most common variants were found in *ABCA4*, with *ABCA4* c.5603A>T (p.Asn1868Ile; n=49), c.5882G>A (p.Gly1961Glu; n=38), and c.5461-10T>C (n=27) most frequently identified. **Supplemental Table 7** summarizes the most frequent variants identified among participants who completed genetic testing. Novel variants are being curated and validated to be reported in a separate manuscript.

**Figure 7.**
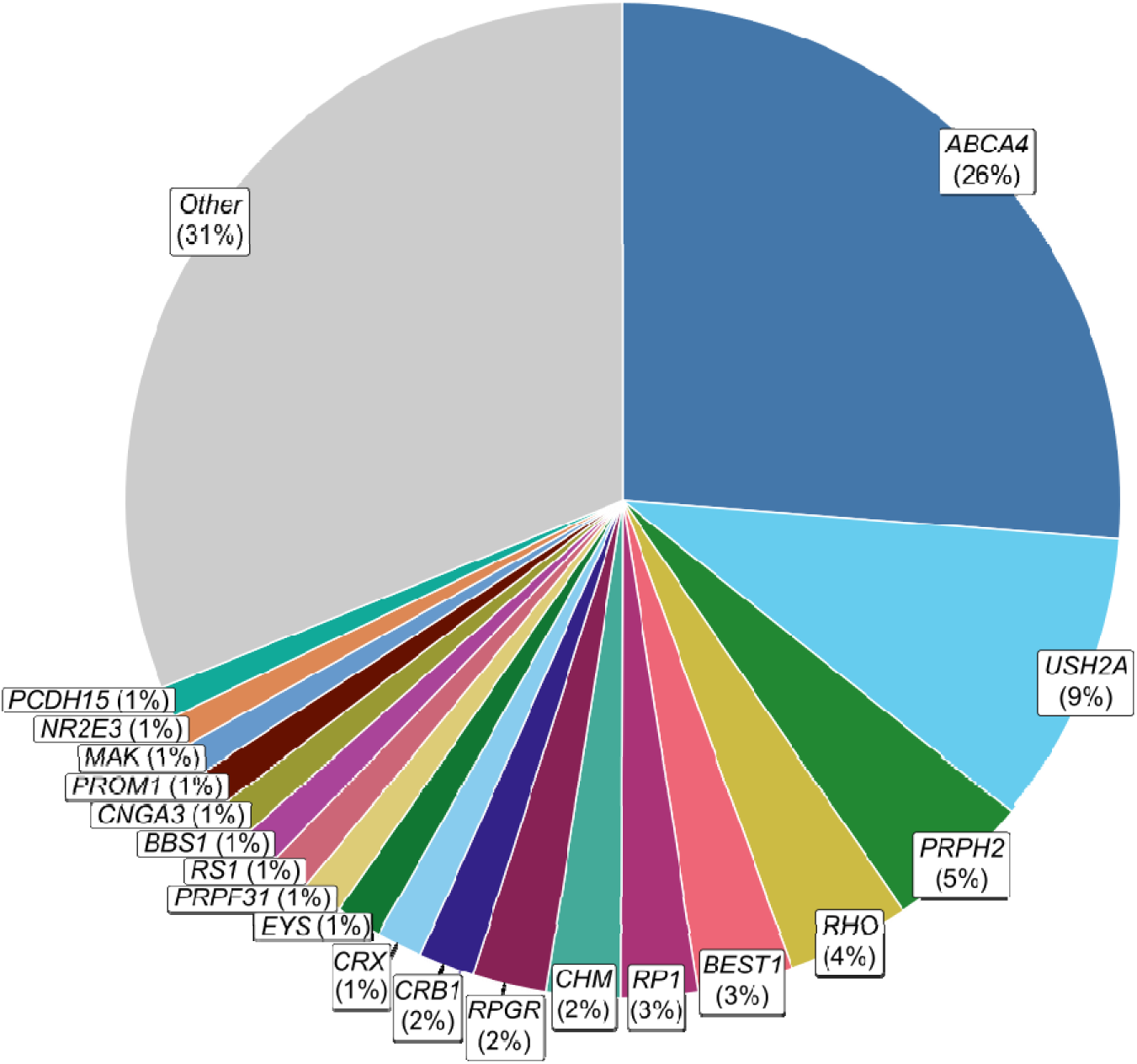
Most common causative genes among participants with likely molecular diagnoses and available variant information. Distribution of 675 gene-level likely molecular diagnoses among 673 participants with likely positive genetic testing results and available variant information. Two participants had dual molecular diagnoses of both *ABCA4* and *PRPH2,* and are counted twice. Genes representing <1% of likely molecular diagnoses are grouped as “Other” (99 genes; comprising 31% of likely molecular diagnoses).

**Table 4.** Cumulative patient counts by most common causative genes.

| Top Genes | Our Cohort | FFB Gene Poll – Global | FFB Gene Poll – U.S. |
| --- | --- | --- | --- |
| **Top 5 genes** | 47.2% | 40.9% | 42% |
| 1 | *ABCA4* (26.2%) | *ABCA4* – 17% | *ABCA4* – 16% |
| 2 | *USH2A* (9.5%) | *USH2A* – 9% | *USH2A* – 9% |
| 3 | *PRPH2* (4.7%) | *RPGR* – 6% | *RPGR* – 7% |
| 4 | *RHO* (4.0%) | *PRPH2* – 5% | *PRPH2* – 5% |
| 5 | *BEST1* (3.1%) | *RHO* – 4% | *RHO* – 5% |
| **Top 20 genes** | 69.4% | 67.9% |  |
| **Top 100 genes** | 96.9% | 94.4% |  |
| **Top 118 genes** | 100.0% |  |  |

### Racial differences in clinical characteristics, genetic testing patterns, and molecular diagnoses

In a subgroup analysis, we compared Black or African American (n=350) and non-Hispanic White (n=902) participants. Age at symptom onset, age at presentation, symptom duration prior to presentation, follow-up duration at Wilmer, and time from presentation to genetic testing did not significantly differ. Black participants had significantly worse BCVA at all time points, after adjusting for multiple comparisons (adjusted p<0.001 for all) (**Table 5**).

**Table 5.** Comparison of clinical and genetic testing characteristics between Black and non-Hispanic White participants with genetic eye disorders. Data presented as median (IQR). Rates of genetic testing completion and molecular diagnosis by race are reported separately in **Supplemental Table 3** and **Table 3**, respectively.

| Parameter | White (N=902) | Black (N=350) | Raw p-value | Adjusted p-value |
| --- | --- | --- | --- | --- |
| Age of symptom onset | 30 (IQR 13-49) | 28 (IQR 13-48) | M-W p=0.514 | 1 |
| Age at presentation | 46 (IQR 29-59) | 44 (IQR 29-55) | M-W p=0.069 | 0.35 |
| Symptom duration to presentation | 5 (IQR 1-20.8) | 6 (IQR 1-16) | M-W p=0.474 | 1 |
| Baseline BCVA, better-seeing eye | 0.19 (IQR 0-0.544) | 0.398 (IQR 0.097-0.796) | M-W p=8.4e-09 | 5.9e-08* |
| Baseline BCVA, worse-seeing eye | 0.398 (IQR 0.097-0.796) | 0.602 (IQR 0.301-1.197) | M-W p=9.4e-11 | 8.5e-10* |
| Follow-up BCVA, better-seeing eye | 0.301 (IQR 0.097-0.875) | 0.602 (IQR 0.176-1.097) | M-W p=1.6e-06 | 9.9e-06* |
| Follow-up BCVA, worse-seeing eye | 0.602 (IQR 0.204-1.176) | 0.903 (IQR 0.398-2) | M-W p=3.9e-10 | 3.2e-09* |
| Duration of follow-up | 6 (IQR 2-12) | 6.5 (IQR 2-13) | M-W p=0.961 | 1 |
| Time from presentation to GT completion (years) | 2 (IQR 0-8) | 2 (IQR 0-9) | M-W p=0.719 | 1 |

\*Adjusted p<0.05 (Holm-Bonferroni correction for multiple comparisons)
Abbreviations: BCVA best-corrected visual acuity (logMAR); GT genetic testing; IQR interquartile range; M-W Mann-Whitney U test

**Table 5.** Comparison of clinical and genetic testing characteristics between Black and non-Hispanic White participants with genetic eye disorders.
| Parameter | White (N=902) | Black (N=350) | Raw p-value | Adjusted p-value |
| --- | --- | --- | --- | --- |
| Age of symptom onset | 30 (IQR 13-49) | 28 (IQR 13-48) | M-W<br>p=0.514 | 1 |
| Age at presentation | 46 (IQR 29-59) | 44 (IQR 29-55) | M-W<br>p=0.069 | 0.35 |
| Symptom duration to presentation | 5 (IQR 1-20.8) | 6 (IQR 1-16) | M-W<br>p=0.474 | 1 |
| Baseline BCVA, better-seeing eye | 0.19 (IQR 0-0.544) | 0.398 (IQR 0.097-0.796) | M-W p=8.4e-09 | 5.9e-08* |
| Baseline BCVA, worse-seeing eye | 0.398 (IQR 0.097-0.796) | 0.602 (IQR 0.301-1.197) | M-W p=9.4e-11 | 8.5e-10* |
| Follow-up BCVA, better-seeing eye | 0.301 (IQR 0.097-0.875) | 0.602 (IQR 0.176-1.097) | M-W p=1.6e-06 | 9.9e-06* |
| Follow-up BCVA, worse-seeing eye | 0.602 (IQR 0.204-1.176) | 0.903 (IQR 0.398-2) | M-W p=3.9e-10 | 3.2e-09* |
| Duration of follow-up | 6 (IQR 2-12) | 6.5 (IQR 2-13) | M-W<br>p=0.961 | 1 |
| Time from presentation to GT completion (years) | 2 (IQR 0-8) | 2 (IQR 0-9) | M-W<br>p=0.719 | 1 |

\*Adjusted p<0.05 (Holm-Bonferroni correction for multiple comparisons)
Abbreviations: BCVA best-corrected visual acuity (logMAR); GT genetic testing; IQR interquartile range; M-W Mann-Whitney U test

Clinical phenotype distributions also differed, with RP more prevalent among Black (37%) than White (31%) participants. Other leading putative diagnoses in White participants were STGD (17%), PD (10%), CD (7%), CRD (6%), and USH (5%), while in Black participants, STGD (17%), CD (8%), CRD (8%), PD (4%), and HON (4%) were next most common (**Supplemental Table 8**). Among participants with a molecular diagnosis, *ABCA4* was the most common gene in both groups (White: 23.9%, Black: 29%). In White participants (n=468), other frequent genes included *USH2A* (11.7%), *PRPH2* (5.3%), *RHO* (5.1%), and *BEST1* (3.9%). In Black participants (n=100), genes that were more frequently detected than in White participants included *SCA7* (6.1%), *CRB1* (4.0%), *RP1* (4.0%), *CNGA3* (3.0%), *PDE6B* (3.0%), and *RPGR* (3.0%) (**Supplemental Table 9**).

Notably, the Other race (n=106) group also exhibited significantly worse BCVA at baseline (adjusted p<0.01 for both eyes) and follow-up (better-seeing eye adjusted p<0.05; worse-seeing eye followed a similar trend though not statistically significant with adjusted p=0.06) compared to the non-Hispanic White group (n=902). Other race participants also had younger age at symptom onset (median 19.5 vs. 30 years; adjusted p<0.001) and presentation (36 vs. 46 years; adjusted p<0.001). The interval from presentation to genetic testing completion was shorter in the Other race group (median 1 vs. 2 years; adjusted p<0.05). Approximately 17% (18/106) of these participants were international referrals, primarily from the Middle East, which may contribute to the shorter duration of follow-up in this group (median 2.5 vs. 6 years; adjusted p<0.001). These differences, particularly the limited follow-up, should be considered when interpreting visual outcomes and may further compound disparities observed in this subgroup (**Supplemental Table 10**).

## Discussion

This study, to our knowledge the largest single-center analysis of GED patients in the U.S. to date, demonstrates that both clinical and demographic factors—including earlier age of symptom onset, Black/African American or Other race, and worse visual function—are key predictors of genetic testing completion and molecular diagnostic yield. These findings highlight actionable and previously underappreciated opportunities for advancing precision medicine and equity in hereditary eye disease care.

Importantly, earlier age of symptom onset emerged as a novel, independent predictor of genetic testing completion. Completion rates were also higher among those with younger age at presentation and longer follow-up, suggesting that disease severity and sustained care engagement may motivate pursuit of a genetic diagnosis.^39,56^ Patients presenting at a younger age are more likely to have severe, early-onset, or syndromic forms of GED, which may prompt clinicians and families to pursue genetic testing for diagnostic clarification, prognostication, and eligibility for emerging gene-based therapies.^48,57,58^ In contrast, Black and Other race participants were substantially less likely to complete genetic testing and receive a molecular diagnosis, even after accounting for clinical variables, and also had consistently worse BCVA compared to non-Hispanic White participants.

Among participants who completed genetic testing, greater symptom duration prior to both presentation and genetic testing completion was associated with higher probability of molecular diagnosis. This finding may reflect the “genetic odyssey” endured by many GED patients, where slow disease progression and diagnostic uncertainty lead to delays in both evaluation and genetic testing.^59^ One possible explanation is that as clinical features evolve over time, they may eventually facilitate more accurate genotype-phenotype correlations and variant interpretation. Alternatively, patients with longer disease duration may have undergone multiple evaluations or testing iterations, increasing the likelihood of an eventual diagnosis. Regardless of mechanism, these delays can postpone access to counseling, trial eligibility, and informed care decisions, particularly among underrepresented groups who may already face systemic barriers to care. Expanding access to genetic testing at the point of diagnosis remains critical.

Diagnostic yield was highest among syndromic or well-characterized monogenic phenotypes (e.g. USH, BEST, STGD) and X-linked recessive conditions (e.g. retinoschisis, choroideremia), moderate in LCA/SEORD, achromatopsia, RP, OCA, and CRD, and lower in heterogeneous or later-onset conditions such as PD, HON, CD, L-ORD, and unspecified macular and retinal dystrophies. These results closely mirror previously reported high yields for USH and STGD (83-94%) and moderate yields for RP (46-67%), reinforcing the need for phenotype-informed counseling and continued gene discovery, particularly in genetically complex or late-onset GEDs.^11,32,35,60–64^ The lower yields observed in some phenotype categories may reflect greater locus heterogeneity, incomplete gene discovery, or phenotypic overlap (e.g. CD/CRD mimicking RP, or PD resembling STGD, other macular dystrophies, or age-related macular degeneration, particularly in older adults).^65,66^ Additionally, younger age, male sex, earlier symptom onset, and worse BCVA were all associated with increased diagnostic yield. We observed a stepwise decline in diagnostic yield with increasing age of onset, consistent with prior studies suggesting that genetically solvable GEDs tend to present earlier and progress more rapidly.^67–69^ The higher yield in males may partly reflect the inclusion of X-linked recessive conditions such as *RPGR-* and *CHM-*associated GEDs. Overall, our diagnostic yield of 62% matches the range reported in previous studies (52-76%).^70–72^

We found that Black and Other race participants were less likely than non-Hispanic White participants to complete genetic testing, receive a molecular diagnosis, or achieve favorable visual outcomes. Our findings validate those of Zhao et al., who reported lower genetic testing completion rates for Black and Other race IRD patients compared to White patients.^73^ Notably, our study is the first to demonstrate that, in addition to Black race, Other race participants also have a significantly lower molecular diagnostic yield.^74^ Importantly, the time from clinical presentation to genetic testing completion was not longer for Black or Other race participants compared to White participants, suggesting comparable workflows once patients are engaged with genetic services, though barriers to initial engagement may persist.

These disparities are likely multifactorial, stemming from differences in healthcare access, trust, health literacy, and the underrepresentation of non-European ancestries in genomic databases.^75,76^ At the population level, similar patterns have been reported: a meta-analysis of over 23,000 IRD patients found higher diagnostic yields in individuals of European ancestry (62.9%) compared to East Asians (54.9%),^75^ and only minimal overlap in pathogenic variants has been observed across ethnic groups.^77^ Such limitations in current genomic resources lead to higher rates of inconclusive results and reduce the generalizability of testing outcomes for diverse populations. Downstream, these disparities likely translate into inequities in access to clinical trials; while race-specific data on GED trials are lacking, underrepresentation of racial minorities in clinical trials is well-documented in other settings.^78,79^ Without deliberate inclusion efforts, emerging therapies risk compounding existing inequities. Addressing these gaps will require a coordinated effort to diversify reference databases, support culturally sensitive counseling models, and ensure equitable inclusion in research.^80^

Compared to international cohorts, our population had an older median age of symptom onset (28 years; reported means 8-20 years)^14,24,31,81^ and baseline evaluation (44 years; reported medians 31-53 years).^14,20,30,35,36,39,82,83^ This difference likely reflects the adult referral base of our clinic and the higher representation of adult-onset phenotypes such as PD, late-onset retinal degeneration (L-ORD), and adult-onset vitelliform macular dystrophy. In contrast, pediatric populations worldwide show higher frequencies of early-onset, syndromic, and recessive GEDs, emphasizing the need to contextualize genetic epidemiology by patient demographics and clinical setting.^57^

The genetic architecture in our cohort paralleled international registries, with *ABCA4*, *USH2A*, *PRPH2*, *RHO*, and *BEST1* accounting for 47.2% of solved cases, while 118 genes were detected overall. Compared with the FFB IRD Gene Poll (where *ABCA4*, *USH2A*, *RPGR*, *PRPH2*, and *RHO* comprised 40.9% of molecular diagnoses), *RPGR* was relatively less common in our cohort, likely reflecting underrepresentation of early-onset X-linked cases in our adult-dominant sample.^41,84^ Most molecular diagnoses involved autosomal recessive inheritance, with homozygosity for the causal variant observed in 21% of recessive cases (14% of all solved cases), a proportion lower than reported in more consanguineous or genetically isolated populations.^13,19^

Importantly, genetic testing frequently revised clinical diagnoses, most notably among macular dystrophies. For example, participants with putative diagnoses of STGD or BEST were found to harbor variants in genes such as *PRPH2* and *PRDM13.* Similarly, PD was linked to *PRPH2* and *ARMS2*, suggesting phenotypic overlap with age-related maculopathies. These findings reinforce the limitations of clinical diagnosis alone and the importance of genetic testing for prognosis, counseling, and treatment planning.

This study has several limitations. First, as a retrospective analysis from a single tertiary referral center, our findings may be influenced by referral and selection bias, with an overrepresentation of participants with more severe disease or greater interest in genetic testing. Second, we did not account for relatedness between participants or systematically collect data on socioeconomic status or insurance coverage, which are factors that likely influence testing uptake. Third, the type and timing of genetic testing varied over the study period, and we did not assess temporal trends in testing adoption, which may have evolved with advances in technology and accessibility. Fourth, although we reviewed VUS against current ClinVar annotations (accessed April 2026) and reclassified genetic tests accordingly, variant interpretation remains an evolving process. Notably, 19 cases with one P/LP and one VUS in an autosomal recessive gene confirmed *in trans* were classified as inconclusive per strict ACMG criteria, despite being managed as clinically solved in our practice given phase and phenotype concordance. These cases may represent true molecular diagnoses pending VUS reclassification, highlighting the gap between strict molecular diagnostic criteria and clinical practice. Fifth, we used the term “likely molecular diagnosis” because phase confirmation was not uniformly obtained for phenotypically concordant participants with two P/LP variants in autosomal recessive genes; this may result in modest overestimation of diagnostic yield for autosomal recessive conditions if a subset of variants reside *in cis* rather than *in trans*. Sixth, genetic counseling involvement was not systematically captured in our database; while genetic counseling is a central part of our practice and genetic results were typically delivered by one of three genetic counselors at our institution (CA, CHS, KSG), we did not assess the impact of genetic counseling on genetic testing completion. Seventh, reasons for not completing genetic testing were not systematically collected and should be examined in future studies. Lastly, an important limitation of this study is that we did not systematically collect data on the specific genetic test performed for each participant, including panel version, gene content, or coverage of challenging regions such as *ORF15* of *RPGR*. As testing technology evolved substantially over the study period, earlier and/or targeted panels may have missed variants now detectable with current methodologies, potentially underestimating diagnostic yield particularly among participants tested in earlier years. This heterogeneity reflects a real-world challenge to IRD practice, where patients present with testing performed across different platforms and time points. At our center, there is an ongoing effort to re-test patients who previously received negative or inconclusive results using updated methodologies; however, this process will take years to complete and broadly reflects the current state of IRD genetic testing practices in the United States.

Our data provide key insights into genetic testing practices, outcomes, and persistent disparities in hereditary eye disease. As the largest single-center characterization of GED patients in the U.S., we highlight how both disease- and participant-level characteristics—including age of onset, race, sex, visual function, and symptom duration—shape genetic testing completion and molecular diagnostic yield. Despite high rates of genetic testing completion and molecular diagnosis at our center, notable disparities in access and outcomes remain, particularly among underrepresented groups. As the therapeutic landscape for GEDs continues to evolve, equitable and systematic integration of genetic testing will be critical for guiding diagnosis, counseling, and therapy eligibility, ensuring that all patients can benefit as GED care advances towards precision medicine.

## Data Availability

All data produced in the present study are available upon reasonable request to the authors

## Abbreviations and Acronyms

ACMG: American College of Medical Genetics and Genomics
AMP: Association for Molecular Pathology
arRP: autosomal recessive retinitis pigmentosa
BCVA: best-corrected visual acuity
BEST: Best disease (e.g. Bestrophinopathy & Best vitelliform macular dystrophy)
CD: cone dystrophy
CI: confidence interval
CLIA: Clinical Laboratory Improvement Amendments
CRD: cone-rod dystrophy
FDA: Food and Drug Administration
FFB: Foundation Fighting Blindness
GED: genetic eye disorder
GEDi: Genetic Eye Disease Center
HIPAA: Health Insurance Portability and Accountability Act
HLE: Hispanic or Latino ethnicity
HON: hereditary optic neuropathy
IRD: inherited retinal disease
IQR: interquartile range
LASSO: least absolute shrinkage and selection operator
LCA/SEORD: Leber Congenital Amaurosis/severe early-onset retinal dystrophy
logMAR: logarithm of the Minimum Angle of Resolution
L-ORD: late-onset retinal degeneration
OCA: oculocutaneous albinism
OR: odds ratio
PD: pattern dystrophy
P/LP: pathogenic or likely pathogenic
RP: retinitis pigmentosa
RPE: retinal pigment epithelium
STGD: Stargardt disease
USH: Usher syndrome
U.S.: United States
VUS: variant of uncertain significance

## Notes

**Conflict of Interest:** No conflicting relationship exists for any author.

### Competing Interest Statement

The authors have declared no competing interest.

### Funding Statement

Financial support included The Joseph Albert Hekimian Fund (MSS), Andreas C. Dracopoulos Professorship (MSS), Andreas C. Dracopoulos and Daniel Finkelstein M.D. Rising Professorship in Ophthalmology (JJD), Wilmer Eye Institute Retina Rising Professorship (IA), and The Stirn Family Fund (JJD, MSS). The sponsor or funding organization had no role in the design or conduct of this research.

### Author Declarations

IRB of Johns Hopkins University gave ethical approval for this work

### Summary of Updates

Stricter exclusion criteria implemented for non-genetic etiologies, participants who had completed genetic testing prior to initial evaluation at our institution, and incomplete demographic or clinical data. Results and figures/tables updated accordingly. This revised paper has been accepted and is currently in press.

