## Supplemental Figure 1 for "Disease and Participant-Related Correlates of Genetic Testing Completion for Hereditary Eye Disorders in a Cohort of Over 1400 Patients"

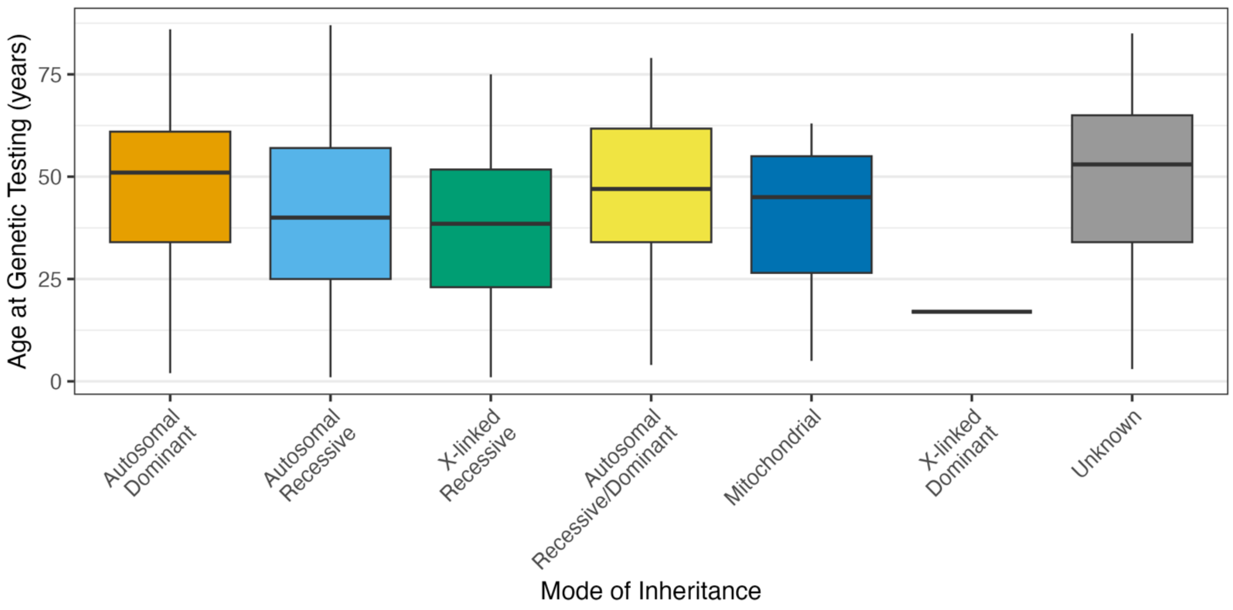


**Supplemental Figure 1. Age at genetic testing completion by mode of inheritance.** Boxplots display median, interquartile range, and range across major inheritance categories (n=1087). 'Unknown' denotes cases where inheritance pattern remained undetermined after testing.
