## Supplemental Table 1 for "Disease and Participant-Related Correlates of Genetic Testing Completion for Hereditary Eye Disorders in a Cohort of Over 1400 Patients"

**Supplemental Table 1.** Demographic, clinical, and genetic characteristics of participants who completed genetic testing prior to presentation (n=111). Data presented as median (IQR) or % (n).

| Parameter | Prior GT Cohort (N=111) |
|---|---|
| **Sex** | |
| Male | 56% (62) |
| Female | 44% (49) |
| **Race** | |
| White | 67% (74) |
| Black or African American | 15% (17) |
| Asian | 13% (14) |
| Other | 5% (6) |
| **Ethnicity** | |
| Hispanic or Latino | 5% (6) |
| Not Hispanic or Latino | 95% (105) |
| **Age** | 42 (IQR 28-58) years |
| **Age of symptom onset** | 21 (IQR 10-41) years |
| **Age at presentation** | 40 (IQR 24-55.5) years |
| **Symptom duration prior to presentation** | 8 (IQR 2-20) years |
| **Age at genetic testing** | 37 (IQR 21.5-51) years |
| **Symptom duration prior to GT completion** | 5 (IQR 0-20.5) years |
| **Duration of follow-up** | 1 (IQR 0-3) years |
| **Baseline BCVA, better-seeing eye** | 0.349 (IQR 0.097-0.679) |
| **Baseline BCVA, worse-seeing eye** | 0.637 (IQR 0.204-1.204) |
| **Follow-up BCVA, better-seeing eye** | 0.301 (IQR 0.097-0.699) |
| **Follow-up BCVA, worse-seeing eye** | 0.602 (IQR 0.176-1.204) |
| **Genetic testing result** | |
| Likely molecular diagnosis | 65% (72) |
| Inconclusive | 23% (25) |
| Negative | 13% (14) |
| **Mode of inheritance (n=72)** | |
| Autosomal Dominant | 15% (11) |
| Autosomal Recessive | 61% (44) |
| X-linked Recessive | 11% (8) |
| Autosomal Recessive/Dominant | 7% (5) |
| Mitochondrial | 4% (3) |

This table describes participants who completed genetic testing prior to initial evaluation at our institution and were excluded from the main analysis. The most common clinical diagnoses were Retinitis Pigmentosa (42.3%), Stargardt disease (17.1%), Achromatopsia (3.6%), Cone Dystrophy (3.6%), Usher syndrome (3.6%). Among participants with likely molecular diagnoses (n=72), the most frequently identified genes were *ABCA4* (n=15), *USH2A* (n=5), *RP1* (n=4), *CRB1* (n=3), and *RPGR* (n=3). Mode of inheritance is reported only for participants with likely molecular diagnoses.

Abbreviations: *BCVA* = best-corrected visual acuity (logMAR); *GT* = genetic testing; *IQR* = interquartile range.
