## Supplemental Table 2 for "Disease and Participant-Related Correlates of Genetic Testing Completion for Hereditary Eye Disorders in a Cohort of Over 1400 Patients"

**Supplemental Table 2.** Putative clinical diagnoses and genetic testing outcomes in the full genetic eye disease cohort (n=1466).

| Putative clinical diagnosis | Age of symptom onset | Age at presentation | Freq | (+)-GT | I-GT | (-)-GT | Untested | Genes identified in participants with likely molecular diagnosis[a] |
|---|---|---|---|---|---|---|---|---|
| Retinitis Pigmentosa | 25 (14-42) | 46 (32-56) | 473 | 44% (207) | 18% (86) | 13% (61) | 25% (119) | *USH2A* (32), *RHO* (25), *RP1* (16), *RPGR* (12), *EYS* (10), *PRPF31* (9), *PRPH2* (8), *MAK* (7), *CRB1* (6), *PDE6B* (6), *NR2E3* (5), *CNGA1* (4), *SNRNP200* (4), *TULP1* (4), *BBS1* (3), *CLN3* (3), *FAM161A* (3), *FLVCR1* (3), *IMPDH1* (3), *PROM1* (3), *RPE65* (3), *CEP290* (2), *IFT140* (2), *NRL* (2), *PRPF8* (2), *ABCA4* (1), *BBS2* (1), *BEST1* (1), *C8ORF37* (1), *CERKL* (1), *CLRN1* (1), *CNGB1* (1), *EXOSC5* (1), *GNPTG* (1), *HGSNAT* (1), *IMPG1* (1), *IMPG2* (1), *MT-TP* (1), *NPHP4* (1), *PDE6A* (1), *PDE6G* (1), *PRPF3* (1), *PRPS1* (1), *RBP3* (1), *RLBP1* (1), *RP2* (1), *SAG* (1), *SCAPER* (1), *SLC52A2* (1), *SPATA7* (1), *TOPORS* (1), *VPS13B/COH1* (1) |
| Stargardt disease | 25 (12-40) | 38 (21-53) | 255 | 71% (182) | 12% (31) | 3% (7) | 14% (35) | *ABCA4* (172), *PRPH2* (6), *ABCA4+PRPH2* (2), *FZD4* (1), *PROM1* (1) |
| Pattern Dystrophy | 60 (47-69) | 60 (50-68) | 106 | 15% (16) | 11% (12) | 18% (19) | 56% (59) | *PRPH2* (12), *IMPG2* (2), *ARMS2* (1), *MT-TS2* (1) |
| Cone Dystrophy | 38 (15-56) | 48 (32-62) | 101 | 18% (18) | 25% (25) | 22% (22) | 36% (36) | *CRB1* (2), *GUCA1A* (2), *GUCY2D* (2), *RDH12* (2), *CACNA2D4* (1), *CNGB3* (1), *CTNNA1* (1), *FAM161A* (1), *KCNV2* (1), *MFSD8/CLN7* (1), *PROM1* (1), *RP1* (1), *RP2* (1), *RPGR* (1) |
| Cone-Rod dystrophy | 30 (18-44) | 44 (31-58) | 99 | 39% (39) | 29% (29) | 12% (12) | 19% (19) | *CRX* (10), *PROM1* (3), *RPGR* (3), *SCA7* (3), *ABCA4* (2), *CACNA1F* (2), *CERKL* (2), *PRPH2* (2), *CEP78* (1), *CLN1* (1), *CNGA3* (1), *CRB1* (1), *GUCY2D* (1), *KIZ* (1), *PRCD* (1), *RET* (1), *RHO* (1), *RPE65* (1), *TTLL5* (1), *USH2A* (1) |
| Usher syndrome | 18 (12-30) | 34 (20-50) | 67 | 84% (56) | 0% (0) | 6% (4) | 10% (7) | *USH2A* (31), *PCDH15* (7), *ADGRV1* (5), *CDH23* (5), *MYO7A* (4), *USH1C* (2), *CLRN1* (1), *GPR98* (1) |
| Hereditary optic neuropathy | 19 (9-39) | 33 (14-42) | 42 | 31% (13) | 19% (8) | 26% (11) | 24% (10) | *OPA1* (4), *ACO2* (3), *MT-ND4* (3), *AFG3L2* (1), *MT-ND1* (1), *MT-ND6* (1) |
| Unspecified macular dystrophy | 38 (25-44) | 42 (31-56) | 39 | 10% (4) | 18% (7) | 28% (11) | 44% (17) | *CDH3* (1), *CRB1* (1), *PRPH2* (1), *TULP1* (1) |
| Best disease (e.g. Bestrophinopathy & Best vitelliform macular dystrophy) | 20 (12-35) | 37 (28-54) | 33 | 64% (21) | 3% (1) | 3% (1) | 30% (10) | *BEST1* (20), *PRDM13* (1) |
| Oculocutaneous Albinism | 0 (0-3) | 22 (6-37) | 25 | 32% (8) | 24% (6) | 4% (1) | 40% (10) | *OCA2* (4), *GPR143* (2), *TYR* (2) |
| L-ORD (late-onset retinal degeneration) | 62.5 (56-70) | 68 (61-77) | 24 | 12% (3) | 17% (4) | 21% (5) | 50% (12) | *C1QTNF5* (3) |
| Achromatopsia | 0.5 (0-9) | 16 (5-37) | 22 | 59% (13) | 9% (2) | 18% (4) | 14% (3) | *CNGA3* (7), *CNGB3* (5), *KCNV2* (1) |
| Retinoschisis | 10 (5-39) | 34 (18-46) | 21 | 48% (10) | 10% (2) | 5% (1) | 38% (8) | *RS1* (9), *OPA1* (1) |
| Unspecified retinal dystrophy | 44.5 (26-55) | 47 (27-64) | 20 | 10% (2) | 40% (8) | 15% (3) | 35% (7) | *MT-ATP6* (1), *NPHP1* (1) |
| Choroideremia | 20.5 (13-41) | 39 (33-57) | 18 | 89% (16) | 6% (1) | 6% (1) | 0% (0) | *CHM* (16) |
| Leber Congenital Amaurosis/Severe early-onset retinal dystrophy (LCA/SEORD) | 0 (0-2) | 9 (3-29) | 17 | 59% (10) | 6% (1) | 18% (3) | 18% (3) | *CEP290* (2), *CRB1* (2), *CABP4* (1), *LCA5* (1), *NPHP4* (1), *RPE65* (1), *RPGRIP1* (1), *SPATA7* (1) |

| Putative clinical diagnosis | Age of symptom onset | Age at presentation | Freq | (+)-GT | I-GT | (-)-GT | Untested | Genes identified in participants with likely molecular diagnosis[a] |
|---|---|---|---|---|---|---|---|---|
| Stickler syndrome | 11 (2-36) | 20 (5-40) | 12 | 50% (6) | 25% (3) | 17% (2) | 8% (1) | *COL2A1* (4), *Col11A1* (1), *LRP2* (1) |
| Mitochondrial retinal dystrophy | 38 (36-42) | 45 (41-49) | 9 | 78% (7) | 11% (1) | 11% (1) | 0% (0) | *MT-TL1* (6), *RHO* (1) |
| Occult macular dystrophy | 25 (13-53) | 48 (32-63) | 9 | 44% (4) | 0% (0) | 33% (3) | 22% (2) | *RP1L1* (4) |
| Pseudoxanthoma elasticum | 53 (38-61) | 64 (44-72) | 9 | 22% (2) | 33% (3) | 0% (0) | 44% (4) | *ABCC6* (2) |
| Bardet-Biedl Syndrome (Laurence-Moon) | 13 (8-24) | 27 (12-34) | 7 | 100% (7) | 0% (0) | 0% (0) | 0% (0) | *BBS1* (5), *BBS2* (1), *BBS4* (1) |
| Congenital Stationary Night Blindness | 10 (6-28) | 12 (6-34) | 7 | 57% (4) | 29% (2) | 0% (0) | 14% (1) | *NYX* (3), *CACNA1F* (1) |
| Adult-onset vitelliform macular dystrophy | 60 (60-67) | 62 (62-69) | 5 | 0% (0) | 20% (1) | 20% (1) | 60% (3) | |
| Blue Cone Monochromacy | 0 (0-3) | 13 (8-15) | 5 | 100% (5) | 0% (0) | 0% (0) | 0% (0) | *OPN1LW/OPN1MW* (3), *OPN1LW* (1), *OPN1MW* (1) |
| Gyrate Atrophy | 5.5 (3-11) | 32 (27-43) | 4 | 50% (2) | 0% (0) | 0% (0) | 50% (2) | *OAT* (2) |
| Hereditary retinal vascular disorder | 32.5 (10-56) | 36 (18-57) | 4 | 0% (0) | 50% (2) | 0% (0) | 50% (2) | |
| Nyctalopia | 46 (30-59) | 44 (27-61) | 4 | 0% (0) | 25% (1) | 25% (1) | 50% (2) | |
| SCA7-related retinal dystrophy (Spinocerebellar ataxia 7) | 19.5 (13-31) | 28 (21-36) | 4 | 75% (3) | 0% (0) | 0% (0) | 25% (1) | *SCA7* (3) |
| Joubert syndrome | 1 (0-2) | 2 (2-3) | 3 | 33% (1) | 0% (0) | 33% (1) | 33% (1) | *C5orf42* (1) |
| LCHAD-related retinitis pigmentosa | 5 (2-10) | 4 (2-17) | 3 | 100% (3) | 0% (0) | 0% (0) | 0% (0) | *HADHA* (3) |
| North Carolina macular dystrophy | 9 (7-39) | 7 (6-44) | 3 | 67% (2) | 0% (0) | 0% (0) | 33% (1) | *PRDM13* (2) |
| Alport syndrome-related retinal dystrophy | 39 (30-48) | 34 (28-40) | 2 | 50% (1) | 0% (0) | 0% (0) | 50% (1) | *COL4A5* (1) |
| Alstrom syndrome | 0 (0-0) | 19 (19-19) | 2 | 100% (2) | 0% (0) | 0% (0) | 0% (0) | *ALMS1* (2) |
| Biette Crystalline Corneoretinal Dystrophy | 36.5 (35-38) | 48 (41-54) | 2 | 100% (2) | 0% (0) | 0% (0) | 0% (0) | *CYP4V2* (2) |
| Central areolar choroidal dystrophy (CACD) | 27 (15-39) | 30 (17-44) | 2 | 50% (1) | 0% (0) | 0% (0) | 50% (1) | *PRPH2* (1) |
| Enhanced S-Cone Syndrome | 12 (8-16) | 18 (17-20) | 2 | 100% (2) | 0% (0) | 0% (0) | 0% (0) | *NR2E3* (2) |
| Batten disease | 5 (5-5) | 5 (5-5) | 1 | 100% (1) | 0% (0) | 0% (0) | 0% (0) | *CLN3* (1) |
| Incontinentia pigmenti | 0 (0-0) | 17 (17-17) | 1 | 100% (1) | 0% (0) | 0% (0) | 0% (0) | *IKBKG* (1) |
| Methylmalonic acidemia-related retinal dystrophy | 0 (0-0) | 21 (21-21) | 1 | 0% (0) | 0% (0) | 0% (0) | 100% (1) | |
| Sorsby fundus dystrophy | 53 (53-53) | 48 (48-48) | 1 | 0% (0) | 100% (1) | 0% (0) | 0% (0) | |
| Von Hippel-Lindau syndrome | 18 (18-18) | 30 (30-30) | 1 | 100% (1) | 0% (0) | 0% (0) | 0% (0) | *VHL* (1) |
| Zellweger spectrum disorder | 0 (0-0) | 1 (1-1) | 1 | 100% (1) | 0% (0) | 0% (0) | 0% (0) | *PEX10* (1) |

| Putative clinical diagnosis | Age of symptom onset | Age at presentation | Freq | (+)-GT | I-GT | (-)-GT | Untested | Genes identified in participants with likely molecular diagnosis[a] |
|---|---|---|---|---|---|---|---|---|

This table summarizes the putative clinical diagnoses assigned at presentation, their frequencies, and corresponding genetic testing (GT) outcomes. For each phenotypic category, the percentage of participants with positive (likely molecular diagnosis), inconclusive, negative, or no genetic testing is reported. Identified causative genes are listed for participants with likely molecular diagnoses in each category.

[a]Note: The number of genes by diagnosis may not match the total number of participants with likely molecular diagnoses, as full genetic testing reports were unavailable for some cases. However, likely molecular diagnosis was confirmed in the medical record for these individuals.
