## Supplemental Table 3 for "Disease and Participant-Related Correlates of Genetic Testing Completion for Hereditary Eye Disorders in a Cohort of Over 1400 Patients"

**Supplemental Table 3.** Comparison of demographic and clinical characteristics by genetic testing status. Data presented as % (n) or median (IQR).

| Parameter | Completed genetic testing (N=1088) | Untested (N=378) | Raw p-value | Adjusted p-value |
|---|---|---|---|---|
| **Sex** | | | $\chi^2$ p=0.903 | 1 |
| Male | 48% (524) | 48% (180) | | |
| Female | 52% (564) | 52% (198) | | |
| **Age** | 50 (IQR 34-66) years | 60 (IQR 44-73) years | M-W p=6.7e-11 | 1.1e-09* |
| **Race** | | | | |
| White | 65% (704) | 52% (198) | $\chi^2$ p=3.3e-05 | 4.3e-04* |
| Black or African American | 20% (219) | 35% (131) | $\chi^2$ p=1.7e-08 | 2.4e-07* |
| Asian | 8% (84) | 4% (17) | $\chi^2$ p=0.08 | 0.8 |
| Native Hawaiian or Other Pacific Islander | <1% (1) | <1% (1) | Fisher's p=0.449 | 1 |
| American Indian or Alaska Native | <1% (3) | 1% (2) | Fisher's p=0.608 | 1 |
| Other | 7% (77) | 8% (29) | $\chi^2$ p=1 | 1 |
| **Ethnicity** | | | $\chi^2$ p=0.662 | 1 |
| Hispanic or Latino | 5% (51) | 4% (15) | | |
| Not Hispanic or Latino | 95% (1037) | 96% (363) | | |
| **Age of symptom onset** | 25 (IQR 10-43) years | 37 (IQR 16-55) years | M-W p=1.1e-10 | 1.7e-09* |
| **Age at presentation** | 41 (IQR 25-55.2) years | 49 (IQR 35-63) years | M-W p=5.4e-11 | 9.2e-10* |
| **Symptom duration prior to presentation** | 6 (IQR 1-19) years | 4 (IQR 1-20) years | M-W p=0.029 | 0.31 |
| **Duration of follow-up** | 6 (IQR 2-12) years | 5 (IQR 1-12) years | M-W p=0.003 | 0.035* |
| **Baseline BCVA, better-seeing eye** | 0.301 (IQR 0.097-0.699) | 0.204 (IQR 0.097-0.602) | M-W p=0.234 | 1 |
| **Baseline BCVA, worse-seeing eye** | 0.477 (IQR 0.176-1) | 0.477 (IQR 0.176-1) | M-W p=0.535 | 1 |
| **Follow-up BCVA, better-seeing eye** | 0.477 (IQR 0.097-1) | 0.398 (IQR 0.097-0.903) | M-W p=0.257 | 1 |
| **Follow-up BCVA, worse-seeing eye** | 0.699 (IQR 0.301-1.301) | 0.699 (IQR 0.204-1.301) | M-W p=0.869 | 1 |

*Adjusted p<0.05 (Holm-Bonferroni correction for multiple comparisons)
Abbreviations: *BCVA* = best-corrected visual acuity (logMAR); *IQR* = interquartile range; *M-W* = Mann-Whitney U test; $\chi^2$ = chi-square test; *Fisher's* = Fisher's exact test.
