## Supplemental Table 4 for "Disease and Participant-Related Correlates of Genetic Testing Completion for Hereditary Eye Disorders in a Cohort of Over 1400 Patients"

**Supplemental Table 4.** Multivariable logistic regression results for factors associated with genetic testing completion. LASSO regression selected race, age of symptom onset, and clinical diagnosis as predictors. For clinical diagnosis, LASSO specifically retained Stargardt disease and pattern dystrophy; all other diagnoses were grouped as 'Other' for comparison against the reference group (retinitis pigmentosa).

| Variable | Level | OR (95% CI) | P-value |
|---|---|---|---|
| Race (ref: White) | American Indian/Alaska Native | 0.42 (0.07-3.36) | 0.36 |
| | Asian | 1.01 (0.59-1.83) | 0.97 |
| | Black or African American | 0.4 (0.3-0.53) | **<0.001** |
| | Native Hawaiian/Pacific Islander | 0.17 (0.01-4.36) | 0.21 |
| | Other race | 0.57 (0.36-0.92) | **0.02** |
| Age of onset, years (ref: 10-19) | 0-9 | 1.53 (1-2.37) | 0.05 |
| | 20-29 | 0.98 (0.63-1.53) | 0.93 |
| | 30-39 | 0.96 (0.62-1.52) | 0.87 |
| | 40-49 | 1 (0.64-1.56) | 0.99 |
| | 50-59 | 0.85 (0.53-1.38) | 0.51 |
| | 60-69 | 0.64 (0.39-1.06) | 0.08 |
| | 70+ | 0.23 (0.12-0.43) | **<0.001** |
| Clinical Diagnosis (ref: Retinitis Pigmentosa) | Other diagnosis | 0.93 (0.69-1.23) | 0.61 |
| | Pattern Dystrophy | 0.35 (0.21-0.57) | **<0.001** |
| | Stargardt disease | 2 (1.32-3.08) | **0.001** |

Predictor selection performed using LASSO regression with 10-fold cross-validation (lambda.1se). Sex, ethnicity, symptom duration, duration of follow-up, and baseline visual acuity were evaluated but not retained by LASSO. Reference categories: White (race), 10-19 years (age of onset), retinitis pigmentosa (clinical diagnosis). Abbreviations: *OR* = odds ratio; *CI* = confidence interval.
