## Supplemental Table 5 for "Disease and Participant-Related Correlates of Genetic Testing Completion for Hereditary Eye Disorders in a Cohort of Over 1400 Patients"

**Supplemental Table 5.** Multivariable logistic regression results for factors associated with molecular diagnostic yield (likely molecular diagnosis vs. inconclusive or negative results). LASSO regression selected sex, race, age of symptom onset, and follow-up BCVA in the better-seeing eye as predictors.

| Variable | Level | OR (95% CI) | P-value |
|---|---|---|---|
| Sex (ref: Female) | Male | 1.37 (1.05-1.78) | **0.02** |
| Race (ref: White) | American Indian/Alaska Native | 0.77 (0.07-16.98) | 0.84 |
| | Asian | 0.68 (0.41-1.13) | 0.13 |
| | Black or African American | 0.37 (0.26-0.51) | **<0.001** |
| | Native Hawaiian/Pacific Islander | 56308.57 (0-NA) | 0.97 |
| | Other race | 0.58 (0.35-0.98) | **0.04** |
| Age of onset, years (ref: 0-9) | 10-19 | 1.03 (0.67-1.59) | 0.88 |
| | 20-29 | 0.8 (0.51-1.28) | 0.35 |
| | 30-39 | 0.7 (0.44-1.13) | 0.14 |
| | 40-49 | 0.52 (0.33-0.82) | **0.005** |
| | 50-59 | 0.2 (0.12-0.33) | **<0.001** |
| | 60-69 | 0.2 (0.11-0.35) | **<0.001** |
| | 70+ | 0.12 (0.04-0.32) | **<0.001** |
| Follow-up BCVA, better eye | Follow-up BCVA, better eye (logMAR) | 1.33 (1.08-1.66) | **0.008** |

Predictor selection performed using LASSO regression with 10-fold cross-validation (lambda.1se). Ethnicity, baseline BCVA (better- and worse-seeing eye), and follow-up BCVA in the worse-seeing eye were evaluated but not retained by LASSO. Reference categories: Female (sex), White (race), 0-9 years (age of onset). Abbreviations: *BCVA* = best-corrected visual acuity (logMAR); *OR* = odds ratio; *CI* = confidence interval.
