## Supplemental Table 6 for "Disease and Participant-Related Correlates of Genetic Testing Completion for Hereditary Eye Disorders in a Cohort of Over 1400 Patients"

**Supplemental Table 6.** Causative genes identified among participants with likely molecular diagnoses.

| Gene | Inheritance Pattern | Frequency | % of participants |
|---|---|---|---|
| *ABCA4* | Autosomal Recessive | 177 | 26.2% |
| *USH2A* | Autosomal Recessive | 64 | 9.5% |
| *PRPH2* | Autosomal Dominant | 32 | 4.7% |
| *RHO* | Autosomal Dominant | 27 | 4% |
| *BEST1* | Autosomal Recessive/Dominant | 21 | 3.1% |
| *RP1* | Autosomal Recessive/Dominant | 17 | 2.5% |
| *CHM* | X-linked Recessive | 16 | 2.4% |
| *RPGR* | X-linked Recessive | 16 | 2.4% |
| *CRB1* | Autosomal Recessive | 12 | 1.8% |
| *CRX* | Autosomal Dominant | 10 | 1.5% |
| *EYS* | Autosomal Recessive | 10 | 1.5% |
| *PRPF31* | Autosomal Dominant | 9 | 1.3% |
| *RS1* | X-linked Recessive | 9 | 1.3% |
| *BBS1* | Autosomal Recessive | 8 | 1.2% |
| *CNGA3* | Autosomal Recessive | 8 | 1.2% |
| *PROM1* | Autosomal Recessive/Dominant | 8 | 1.2% |
| *MAK* | Autosomal Recessive | 7 | 1% |
| *NR2E3* | Autosomal Recessive | 7 | 1% |
| *PCDH15* | Autosomal Recessive | 7 | 1% |
| *CNGB3* | Autosomal Recessive | 6 | 0.9% |
| *MT-TL1* | Mitochondrial | 6 | 0.9% |
| *PDE6B* | Autosomal Recessive | 6 | 0.9% |
| *SCA7* | Autosomal Dominant | 6 | 0.9% |
| *ADGRV1* | Autosomal Recessive | 5 | 0.7% |
| *CDH23* | Autosomal Recessive | 5 | 0.7% |
| *OPA1* | Autosomal Dominant | 5 | 0.7% |
| *RPE65* | Autosomal Recessive | 5 | 0.7% |
| *TULP1* | Autosomal Recessive | 5 | 0.7% |

| Gene | Inheritance Pattern | Frequency | % of participants |
|---|---|---|---|
| *CEP290* | Autosomal Recessive | 4 | 0.6% |
| *CLN3* | Autosomal Recessive | 4 | 0.6% |
| *CNGA1* | Autosomal Recessive | 4 | 0.6% |
| *COL2A1* | Autosomal Dominant | 4 | 0.6% |
| *FAM161A* | Autosomal Recessive | 4 | 0.6% |
| *MYO7A* | Autosomal Recessive | 4 | 0.6% |
| *OCA2* | Autosomal Recessive | 4 | 0.6% |
| *RP1L1* | Autosomal Dominant | 4 | 0.6% |
| *SNRNP200* | Autosomal Dominant | 4 | 0.6% |
| *ACO2* | Unknown after genetic testing | 3 | 0.4% |
| *C1QTNF5* | Autosomal Dominant | 3 | 0.4% |
| *CACNA1F* | X-linked Recessive | 3 | 0.4% |
| *CERKL* | Autosomal Recessive | 3 | 0.4% |
| *FLVCR1* | Autosomal Recessive | 3 | 0.4% |
| *GUCY2D* | Autosomal Dominant | 3 | 0.4% |
| *HADHA* | Autosomal Recessive | 3 | 0.4% |
| *IMPDH1* | Autosomal Dominant | 3 | 0.4% |
| *IMPG2* | Autosomal Recessive | 3 | 0.4% |
| *MT-ND4* | Mitochondrial | 3 | 0.4% |
| *NYX* | X-linked Recessive | 3 | 0.4% |
| *OPN1LW/OPN1 MW* | X-linked Recessive | 3 | 0.4% |
| *PRDM13* | Autosomal Dominant | 3 | 0.4% |
| *ABCC6* | Autosomal Recessive | 2 | 0.3% |
| *ALMS1* | Autosomal Recessive | 2 | 0.3% |
| *BBS2* | Autosomal Recessive | 2 | 0.3% |
| *CLRN1* | Autosomal Recessive | 2 | 0.3% |
| *CYP4V2* | Autosomal Recessive | 2 | 0.3% |
| *GPR143* | X-linked Recessive | 2 | 0.3% |
| *GUCA1A* | Autosomal Dominant | 2 | 0.3% |

| Gene | Inheritance Pattern | Frequency | % of participants |
|---|---|---|---|
| *IFT140* | Autosomal Recessive | 2 | 0.3% |
| *KCNV2* | Autosomal Recessive | 2 | 0.3% |
| *NPHP4* | Autosomal Recessive | 2 | 0.3% |
| *NRL* | Autosomal Recessive | 2 | 0.3% |
| *OAT* | Autosomal Recessive | 2 | 0.3% |
| *PRPF8* | Autosomal Dominant | 2 | 0.3% |
| *RDH12* | Autosomal Recessive | 2 | 0.3% |
| *RP2* | X-linked Recessive | 2 | 0.3% |
| *SPATA7* | Autosomal Recessive | 2 | 0.3% |
| *TYR* | Autosomal Recessive | 2 | 0.3% |
| *USH1C* | Autosomal Recessive | 2 | 0.3% |
| *AFG3L2* | Autosomal Dominant | 1 | 0.1% |
| *ARMS2* | Autosomal Recessive | 1 | 0.1% |
| *BBS4* | Autosomal Recessive | 1 | 0.1% |
| *C5orf42* | Autosomal Recessive | 1 | 0.1% |
| *C8ORF37* | Autosomal Recessive | 1 | 0.1% |
| *CABP4* | Autosomal Recessive | 1 | 0.1% |
| *CACNA2D4* | Autosomal Recessive | 1 | 0.1% |
| *CDH3* | Autosomal Recessive | 1 | 0.1% |
| *CEP78* | Autosomal Recessive | 1 | 0.1% |
| *CLN1* | Autosomal Recessive | 1 | 0.1% |
| *CNGB1* | Autosomal Recessive | 1 | 0.1% |
| *COL4A5* | X-linked Recessive | 1 | 0.1% |
| *CTNNA1* | Autosomal Dominant | 1 | 0.1% |
| *Col11A1* | Autosomal Dominant | 1 | 0.1% |
| *EXOSC5* | Autosomal Recessive | 1 | 0.1% |
| *FZD4* | Unknown after genetic testing | 1 | 0.1% |
| *GNPTG* | Autosomal Recessive | 1 | 0.1% |
| *GPR98* | Autosomal Recessive | 1 | 0.1% |
| *HGSNAT* | Autosomal Recessive | 1 | 0.1% |

| Gene | Inheritance Pattern | Frequency | % of participants |
|---|---|---|---|
| *IKBKG* | X-linked Dominant | 1 | 0.1% |
| *IMPG1* | Autosomal Recessive | 1 | 0.1% |
| *KIZ* | Autosomal Recessive | 1 | 0.1% |
| *LCA5* | Autosomal Recessive | 1 | 0.1% |
| *LRP2* | Autosomal Recessive | 1 | 0.1% |
| *MFSD8/CLN7* | Autosomal Recessive | 1 | 0.1% |
| *MT-ATP6* | Mitochondrial | 1 | 0.1% |
| *MT-ND1* | Mitochondrial | 1 | 0.1% |
| *MT-ND6* | Mitochondrial | 1 | 0.1% |
| *MT-TP* | Mitochondrial | 1 | 0.1% |
| *MT-TS2* | Mitochondrial | 1 | 0.1% |
| *NPHP1* | Autosomal Recessive | 1 | 0.1% |
| *OPN1LW* | X-linked Recessive | 1 | 0.1% |
| *OPN1MW* | X-linked Recessive | 1 | 0.1% |
| *PDE6A* | Autosomal Recessive | 1 | 0.1% |
| *PDE6G* | Autosomal Recessive | 1 | 0.1% |
| *PEX10* | Autosomal Recessive | 1 | 0.1% |
| *PRCD* | Autosomal Recessive | 1 | 0.1% |
| *PRPF3* | Autosomal Dominant | 1 | 0.1% |
| *PRPS1* | X-linked Recessive | 1 | 0.1% |
| *RBP3* | Autosomal Recessive | 1 | 0.1% |
| *RET* | Autosomal Dominant | 1 | 0.1% |
| *RLBP1* | Autosomal Recessive | 1 | 0.1% |
| *RPGRIP1* | Autosomal Recessive | 1 | 0.1% |
| *SAG* | Autosomal Dominant | 1 | 0.1% |
| *SCAPER* | Autosomal Recessive | 1 | 0.1% |
| *SLC52A2* | Autosomal Recessive | 1 | 0.1% |
| *TOPORS* | Autosomal Dominant | 1 | 0.1% |
| *TTLL5* | Autosomal Recessive | 1 | 0.1% |
| *VHL* | Autosomal Dominant | 1 | 0.1% |

| Gene | Inheritance Pattern | Frequency | % of participants |
|---|---|---|---|
| *VPS13B/COH1* | Autosomal Recessive | 1 | 0.1% |

This table lists all 118 unique genes identified among 673 participants with likely molecular diagnoses, representing a total of 675 gene-level diagnoses. 2 participants had dual molecular diagnoses with *ABCA4* and *PRPH2*. Although 676 participants had positive results (likely molecular diagnoses), 3 were excluded due to unavailable gene-level data. Frequencies represent the number of participants in whom each gene was implicated; participants with dual diagnoses are included in both relevant gene rows.
