## Supplemental Table 7 for "Disease and Participant-Related Correlates of Genetic Testing Completion for Hereditary Eye Disorders in a Cohort of Over 1400 Patients"

**Supplemental Table 7.** Frequent variants observed in five or more participants.

| Gene | Codon | Protein | Freq | ClinVar ID | ClinVar Pathogenicity Classification |
|---|---|---|---|---|---|
| *ABCA4* | c.5603A>T | p.Asn1868Ile | 49 | 99390 | Conflicting classifications of pathogenicity [Pathogenic(1); Likely pathogenic(2); Established risk allele(2); Uncertain significance(7); Benign(3); Likely benign(6)] |
| *ABCA4* | c.5882G>A | p.Gly1961Glu | 38 | 7888 | Pathogenic |
| *ABCA4* | c.5461-10T>C | | 27 | 92870 | Pathogenic/Likely pathogenic |
| *ABCA4* | c.6320G>A | p.Arg2107His | 21 | 99448 | Conflicting classifications of pathogenicity [Pathogenic(10); Likely pathogenic(8); Uncertain significance(1)] |
| *ABCA4* | c.2588G>C | p.Gly863Ala | 19 | 7879 | Pathogenic |
| *BBS1* | c.1169T>G | p.Met390Arg | 14 | 12143 | Pathogenic/Likely pathogenic |
| *USH2A* | c.2299del | p.Glu767SerfsTer21 | 14 | 2351 | Pathogenic |
| *USH2A* | c.2276G>T | p.Cys759Phe | 13 | 2356 | Pathogenic |
| *ABCA4* | c.3113C>T | p.Ala1038Val | 12 | 7894 | Pathogenic/Likely pathogenic |
| *ABCA4* | c.4139C>T | p.Pro1380Leu | 11 | 7904 | Pathogenic |
| *ABCA4* | c.1622T>C | p.Leu541Pro | 9 | 99067 | Conflicting classifications of pathogenicity [Pathogenic(21); Likely pathogenic(1); Uncertain significance(1)] |
| *ABCA4* | c.6079C>T | p.Leu2027Phe | 9 | 7882 | Pathogenic/Likely pathogenic |
| *ABCA4* | c.4253+43G>A | | 8 | 99265 | Conflicting classifications of pathogenicity [Pathogenic(2); Likely pathogenic(3); Uncertain significance(2)] |
| *ABCA4* | c.2966T>C | p.Val989Ala | 7 | 99180 | Conflicting classifications of pathogenicity [Pathogenic(6); Likely pathogenic(1); Uncertain significance(1)] |
| *CNGB3* | c.1148del | p.Thr383IlefsTer13 | 7 | 5225 | Pathogenic/Likely pathogenic |
| *CRB1* | c.2506C>A | p.Pro836Thr | 7 | 372352 | Pathogenic/Likely pathogenic |
| *FAM161A* | c.1355_1356del | p.Thr452SerfsTer3 | 7 | 37 | Pathogenic |
| *NR2E3* | c.119-2A>C | | 7 | 191059 | Pathogenic/Likely pathogenic |
| *ABCA4* | c.618C>G | p.Ser206Arg | 6 | 99434 | Conflicting classifications of pathogenicity [Pathogenic(1); Uncertain significance(5); Benign(2)] |

| Gene | Codon | Protein | Freq | ClinVar ID | ClinVar Pathogenicity Classification |
|---|---|---|---|---|---|
| *HGSNAT* | c.1843G>A | p.Ala615Thr | 6 | 208816 | Conflicting classifications of pathogenicity [Pathogenic(2); Likely pathogenic(2); Uncertain significance(6); Benign(2); Likely benign(5)] |
| *MAK* | c.1297_1298insAlu | p.Lys433insAlu | 6 | 2579195 | Pathogenic |
| *ABCA4* | c.5714+5G>A | | 5 | 99403 | Pathogenic |
| *ABCA4* | c.6148G>C | p.Val2050Leu | 5 | 7884 | Likely benign |
| *ABCA4* | c.769-784C>T | | 5 | 2578578 | Conflicting classifications of pathogenicity [Pathogenic(2); Likely pathogenic(1); Uncertain significance(3)] |
| *CNGA3* | c.967G>C | p.Ala323Pro | 5 | 284032 | Conflicting classifications of pathogenicity [Pathogenic(6); Likely pathogenic(1); Uncertain significance(1)] |
| *PRPH2* | c.422A>G | p.Tyr141Cys | 5 | 98666 | Pathogenic/Likely pathogenic |
| *RHO* | c.68C>A | p.Pro23His | 5 | 13013 | Pathogenic |
| *USH2A* | c.7595-3C>G | | 5 | 197447 | Pathogenic/Likely pathogenic |

This table lists all 29 variants observed in ≥5 participants (representing 13.9% of all variant observations in participants who completed genetic testing). These frequent variants span 12 genes, with *ABCA4* accounting for 16 variants. Gene name, cDNA change (codon), protein change, observed frequency (Freq), ClinVar variation ID, and ClinVar pathogenicity classification (as of January 2026) are provided.
