## Supplemental Table 8 for "Disease and Participant-Related Correlates of Genetic Testing Completion for Hereditary Eye Disorders in a Cohort of Over 1400 Patients"

**Supplemental Table 8.** Comparison of putative clinical diagnoses between Black and White participants.

| Putative clinical diagnosis | White (n) | White (%; n=902) | Black (n) | Black (%; n=350) |
|---|---|---|---|---|
| Retinitis Pigmentosa | 278 | 30.82% | 130 | 37.14% |
| Stargardt disease | 155 | 17.18% | 60 | 17.14% |
| Pattern Dystrophy | 87 | 9.65% | 15 | 4.29% |
| Cone Dystrophy | 60 | 6.65% | 29 | 8.29% |
| Cone-Rod dystrophy | 56 | 6.21% | 28 | 8% |
| Usher syndrome | 49 | 5.43% | 10 | 2.86% |
| Unspecified macular dystrophy | 24 | 2.66% | 9 | 2.57% |
| Best disease (e.g. Bestrophinopathy & Best vitelliform macular dystrophy) | 23 | 2.55% | 4 | 1.14% |
| L-ORD (late-onset retinal degeneration) | 18 | 2% | 3 | 0.86% |
| Oculocutaneous Albinism | 18 | 2% | 5 | 1.43% |
| Hereditary optic neuropathy | 15 | 1.66% | 15 | 4.29% |
| Retinoschisis | 14 | 1.55% | 6 | 1.71% |
| Choroideremia | 13 | 1.44% | 3 | 0.86% |
| Achromatopsia | 10 | 1.11% | 6 | 1.71% |
| Unspecified retinal dystrophy | 10 | 1.11% | 5 | 1.43% |
| Stickler syndrome | 9 | 1% | 3 | 0.86% |
| Mitochondrial retinal dystrophy | 8 | 0.89% | 1 | 0.29% |
| LCA/SEORD | 7 | 0.78% | 3 | 0.86% |
| Pseudoxanthoma elasticum | 6 | 0.67% | 1 | 0.29% |
| Occult macular dystrophy | 5 | 0.55% | 3 | 0.86% |
| Adult-onset vitelliform macular dystrophy | 4 | 0.44% | 0 | 0% |
| Bardet-Biedl Syndrome (Laurence-Moon) | 4 | 0.44% | 1 | 0.29% |
| Blue Cone Monochromacy | 4 | 0.44% | 1 | 0.29% |
| Congenital Stationary Night Blindness | 4 | 0.44% | 1 | 0.29% |
| LCHAD-related retinitis pigmentosa | 3 | 0.33% | 0 | 0% |
| Nyctalopia | 3 | 0.33% | 1 | 0.29% |
| Enhanced S-Cone Syndrome | 2 | 0.22% | 0 | 0% |
| Gyrate Atrophy | 2 | 0.22% | 0 | 0% |
| Hereditary retinal vascular disorder | 2 | 0.22% | 1 | 0.29% |
| North Carolina macular dystrophy | 2 | 0.22% | 1 | 0.29% |
| Alport syndrome-related retinal dystrophy | 1 | 0.11% | 1 | 0.29% |
| Batten disease | 1 | 0.11% | 0 | 0% |
| Biette Crystalline Corneoretinal Dystrophy | 1 | 0.11% | 0 | 0% |
| Central areolar choroidal dystrophy (CACD) | 1 | 0.11% | 0 | 0% |
| Methylmalonic acidemia-related retinal dystrophy | 1 | 0.11% | 0 | 0% |
| Sorsby fundus dystrophy | 1 | 0.11% | 0 | 0% |
| Von Hippel-Lindau syndrome | 1 | 0.11% | 0 | 0% |
| SCA7-related retinal dystrophy (Spinocerebellar ataxia 7) | 0 | 0% | 4 | 1.14% |

Frequencies and percentages are column-based within each racial group.
