## Supplemental Table 9 for "Disease and Participant-Related Correlates of Genetic Testing Completion for Hereditary Eye Disorders in a Cohort of Over 1400 Patients"

**Supplemental Table 9.** Comparison of gene frequencies among White (n=468) and Black (n=100) participants with likely molecular diagnoses. Only genes identified in at least one participant are shown. Percentages are calculated within racial groups.

| Gene Identified | Frequency (White) | Percent (White; n=468) | Frequency (Black) | Percent (Black; n=100) |
|---|---|---|---|---|
| *ABCA4* | 112 | 23.93% | 29 | 29% |
| *USH2A* | 55 | 11.75% | 3 | 3% |
| *PRPH2* | 25 | 5.34% | 4 | 4% |
| *RHO* | 24 | 5.13% | 2 | 2% |
| *BEST1* | 18 | 3.85% | 1 | 1% |
| *CHM* | 12 | 2.56% | 3 | 3% |
| *RP1* | 10 | 2.14% | 4 | 4% |
| *RPGR* | 10 | 2.14% | 3 | 3% |
| *PRPF31* | 8 | 1.71% | | |
| *CRX* | 7 | 1.5% | 1 | 1% |
| *MAK* | 7 | 1.5% | | |
| *NR2E3* | 7 | 1.5% | | |
| *RS1* | 7 | 1.5% | 2 | 2% |
| *BBS1* | 6 | 1.28% | 1 | 1% |
| *MT-TL1* | 6 | 1.28% | | |
| *PCDH15* | 6 | 1.28% | 1 | 1% |
| *CDH23* | 4 | 0.85% | 1 | 1% |
| *CNGA1* | 4 | 0.85% | | |
| *CNGB3* | 4 | 0.85% | 1 | 1% |
| *CRB1* | 4 | 0.85% | 4 | 4% |
| *SNRNP200* | 4 | 0.85% | | |
| *TULP1* | 4 | 0.85% | 1 | 1% |
| *ADGRV1* | 3 | 0.64% | | |
| *C1QTNF5* | 3 | 0.64% | | |
| *CERKL* | 3 | 0.64% | | |
| *EYS* | 3 | 0.64% | 2 | 2% |
| *FAM161A* | 3 | 0.64% | | |
| *FLVCR1* | 3 | 0.64% | | |
| *HADHA* | 3 | 0.64% | | |
| *IMPDH1* | 3 | 0.64% | | |
| *IMPG2* | 3 | 0.64% | | |
| *NYX* | 3 | 0.64% | | |
| *OPA1* | 3 | 0.64% | 1 | 1% |
| *PDE6B* | 3 | 0.64% | 3 | 3% |
| *PRDM13* | 3 | 0.64% | | |
| *PROM1* | 3 | 0.64% | 2 | 2% |
| *RPE65* | 3 | 0.64% | 2 | 2% |
| *ABCA4+PRPH2* | 2 | 0.43% | | |
| *ABCC6* | 2 | 0.43% | | |
| *CEP290* | 2 | 0.43% | 1 | 1% |
| *CLN3* | 2 | 0.43% | 1 | 1% |
| *CNGA3* | 2 | 0.43% | 3 | 3% |
| *COL2A1* | 2 | 0.43% | 2 | 2% |
| *GPR143* | 2 | 0.43% | | |
| *GUCA1A* | 2 | 0.43% | | |
| *IFT140* | 2 | 0.43% | | |
| *KCNV2* | 2 | 0.43% | | |
| *MT-ND4* | 2 | 0.43% | 1 | 1% |

| Gene Identified | Frequency (White) | Percent (White; n=468) | Frequency (Black) | Percent (Black; n=100) |
|---|---|---|---|---|
| *MYO7A* | 2 | 0.43% | | |
| *NRL* | 2 | 0.43% | | |
| *OPN1LW/OPN1MW* | 2 | 0.43% | 1 | 1% |
| *RP1L1* | 2 | 0.43% | 2 | 2% |
| *RP2* | 2 | 0.43% | | |
| *TYR* | 2 | 0.43% | | |
| *USH1C* | 2 | 0.43% | | |
| *AFG3L2* | 1 | 0.21% | | |
| *ARMS2* | 1 | 0.21% | | |
| *BBS2* | 1 | 0.21% | | |
| *C8ORF37* | 1 | 0.21% | | |
| *CACNA2D4* | 1 | 0.21% | | |
| *CEP78* | 1 | 0.21% | | |
| *CLN1* | 1 | 0.21% | | |
| *CLRN1* | 1 | 0.21% | | |
| *COL4A5* | 1 | 0.21% | | |
| *CTNNA1* | 1 | 0.21% | | |
| *CYP4V2* | 1 | 0.21% | | |
| *Col11A1* | 1 | 0.21% | | |
| *EXOSC5* | 1 | 0.21% | | |
| *FZD4* | 1 | 0.21% | | |
| *GNPTG* | 1 | 0.21% | | |
| *GPR98* | 1 | 0.21% | | |
| *GUCY2D* | 1 | 0.21% | 2 | 2% |
| *HGSNAT* | 1 | 0.21% | | |
| *KIZ* | 1 | 0.21% | | |
| *LRP2* | 1 | 0.21% | | |
| *MFSD8/CLN7* | 1 | 0.21% | | |
| *MT-ATP6* | 1 | 0.21% | | |
| *MT-ND6* | 1 | 0.21% | | |
| *MT-TS2* | 1 | 0.21% | | |
| *NPHP1* | 1 | 0.21% | | |
| *OAT* | 1 | 0.21% | | |
| *OCA2* | 1 | 0.21% | 2 | 2% |
| *OPN1LW* | 1 | 0.21% | | |
| *OPN1MW* | 1 | 0.21% | | |
| *PDE6A* | 1 | 0.21% | | |
| *PDE6G* | 1 | 0.21% | | |
| *PRCD* | 1 | 0.21% | | |
| *PRPF3* | 1 | 0.21% | | |
| *PRPF8* | 1 | 0.21% | 1 | 1% |
| *PRPS1* | 1 | 0.21% | | |
| *RBP3* | 1 | 0.21% | | |
| *RET* | 1 | 0.21% | | |
| *RPGRIP1* | 1 | 0.21% | | |
| *SAG* | 1 | 0.21% | | |
| *SLC52A2* | 1 | 0.21% | | |
| *TOPORS* | 1 | 0.21% | | |
| *VHL* | 1 | 0.21% | | |
| *VPS13B/COH1* | 1 | 0.21% | | |
| *ACO2* | | | 2 | 2% |
| *BBS4* | | | 1 | 1% |
| *CACNA1F* | | | 1 | 1% |

| Gene Identified | Frequency (White) | Percent (White; n=468) | Frequency (Black) | Percent (Black; n=100) |
|---|---|---|---|---|
| *IMPG1* | | | 1 | 1% |
| *MT-ND1* | | | 1 | 1% |
| *MT-TP* | | | 1 | 1% |
| *SCA7* | | | 6 | 6% |
