## Supplemental Table 10 for "Disease and Participant-Related Correlates of Genetic Testing Completion for Hereditary Eye Disorders in a Cohort of Over 1400 Patients"

**Supplemental Table 10.** Comparison of clinical and genetic testing characteristics between non-Hispanic White and Other race participants with genetic eye disorders. Data presented as median (IQR). Rates of genetic testing completion and likely molecular diagnosis by race are reported separately in **Supplemental Table 3** and **Table 3**, respectively.

| Parameter | White (N=902) | Other (N=106) | Raw p-value | Adjusted p-value |
|---|---|---|---|---|
| Age of symptom onset | 30 (IQR 13-49) | 19.5 (IQR 6-34.8) | M-W p=6.0e-05 | 4.2e-04* |
| Age at presentation | 46 (IQR 29-59) | 36 (IQR 20.2-45.8) | M-W p=3.4e-06 | 2.7e-05* |
| Symptom duration to presentation | 5 (IQR 1-20.8) | 8 (IQR 2-18) | M-W p=0.358 | 0.36 |
| Baseline BCVA, better-seeing eye | 0.19 (IQR 0-0.544) | 0.398 (IQR 0.097-0.856) | M-W p=0.001 | 0.008* |
| Baseline BCVA, worse-seeing eye | 0.398 (IQR 0.097-0.796) | 0.544 (IQR 0.301-1) | M-W p=0.001 | 0.008* |
| Follow-up BCVA, better-seeing eye | 0.301 (IQR 0.097-0.875) | 0.61 (IQR 0.176-1) | M-W p=0.011 | 0.034* |
| Follow-up BCVA, worse-seeing eye | 0.602 (IQR 0.204-1.176) | 0.799 (IQR 0.398-1.277) | M-W p=0.03 | 0.06 |
| Duration of follow-up | 6 (IQR 2-12) | 2.5 (IQR 1-7) | M-W p=2.5e-08 | 2.3e-07* |
| Time from presentation to GT completion (years) | 2 (IQR 0-8) | 1 (IQR 0-3) | M-W p=0.003 | 0.012* |

*Adjusted p<0.05 (Holm-Bonferroni correction for multiple comparisons)
Abbreviations: BCVA best-corrected visual acuity (logMAR); GT genetic testing; IQR interquartile range; M-W Mann-Whitney U test
